# PA-FGRS is a novel estimator of pedigree-based genetic liability that complements genotype-based inferences into the genetic architecture of major depressive disorder

**DOI:** 10.1101/2023.06.23.23291611

**Authors:** Morten Dybdahl Krebs, Kajsa-Lotta Georgii Hellberg, Mischa Lundberg, Vivek Appadurai, Henrik Ohlsson, Emil Pedersen, Jette Steinbach, Jamie Matthews, Sonja LaBianca, Xabier Calle, Joeri J. Meijsen, iPSYCH Study Consortium, Andrés Ingason, Alfonso Buil, Bjarni J. Vilhjálmsson, Jonathan Flint, Silviu-Alin Bacanu, Na Cai, Andy Dahl, Noah Zaitlen, Thomas Werge, Kenneth S. Kendler, Andrew J. Schork

## Abstract

Large biobank samples provide an opportunity to integrate broad phenotyping, familial records, and molecular genetics data to study complex traits and diseases. We introduce Pearson-Aitken Family Genetic Risk Scores (PA-FGRS), a new method for estimating disease liability from patterns of diagnoses in extended, age-censored genealogical records. We then apply the method to study a paradigmatic complex disorder, Major Depressive Disorder (MDD), using the iPSYCH2015 case-cohort study of 30,949 MDD cases, 39,655 random population controls, and more than 2 million relatives. We show that combining PA-FGRS liabilities estimated from family records with molecular genotypes of probands improves the three lines of inquiry. Incorporating PA-FGRS liabilities improves classification of MDD over and above polygenic scores, identifies robust genetic contributions to clinical heterogeneity in MDD associated with comorbidity, recurrence, and severity, and can improve the power of genome-wide association studies (GWAS). Our method is flexible and easy to use and our study approaches are generalizable to other data sets and other complex traits and diseases.

## Introduction

The analysis of large biobanks (e.g., BioBank Japan^1^, deCODE genetics^2^, iPSYCH^3, 4^, UKBiobank^5^, etc.) is omnipresent in complex disorder genetics research. These resources provide opportunities to combine large samples, molecular data, diverse phenotypes, and familial phenotypes. Leveraging familial phenotypes to estimate disease liability in large biobanks has applications for improving power of genome-wide association studies (GWAS)^6, 7^, making classifications and predictions^8–10^, and offering better descriptions of underlying causes of disease and heterogeneity^11, 12^. Combining familial and molecular data for these questions may be especially relevant for paradigmatic complex disorders, such as Major Depressive Disorder (MDD), a leading cause of disability world-wide. Such disorders are marked by complex, multifactorial, highly polygenic etiologies that limit the power of molecular genetic investigations^13, 14^, meaning improved approaches are needed. However, it is not clear how best to combine familial phenotypes and genotype data. Existing methods can not fully accommodate all biobanks, including the largest for psychiatric genetics, the iPSYCH2015 case-cohort study, due to complex, age-censored, extended genealogies. Previous applications have focused on one use-case (e.g., GWAS *or* prediction) limiting the picture of generalizability to other questions. Here, we set out to develop a method that is applicable to any biobank and demonstrate, by studying the genetic basis of MDD, that it can improve multiple paradigms applied in molecular genetic studies of complex disorders.

Currently, methods that transform patterns of diagnoses in genealogies to continuous liability^15, 16^ scores in each relative are limited. LT-FH^7^ and LT-FH++^17^ use similar resampling approaches to estimate posterior mean liabilities of relatives, but only consider first degree relatives. This excludes information from more distant relatives and could confound estimates more strongly with familial environment. Both were applied only in the context of improving GWAS. So et al.^18^ developed a method based on the Pearson-Aitken (PA) selection formula^19^, that is an analytical procedure for calculating liability from phenotypes in arbitrarily structured genealogies, but assume each relative has been followed for their entire life (i.e., is fully observed). A flexible, resampling-based extension of this model was proposed, but is computationally prohibitive at scale^20^. These approaches have had a focus on trait predictions. Family Genetic Risk Scores (FGRS)^21^ are kinship weighted sums of diagnoses of relatives with corrections for familial environment, censoring, and other covariates. FGRS accommodates extended genealogies and censored records, but is not based on a well-described model and does not account for kinship among relatives of probands. FGRS has been applied to describe genetic heterogeneity within and across disorders. Current methods estimating individual liability from genealogies are limited and have been applied narrowly.

We introduce a new method, Pearson-Aitken Family Genetic Risk Scores (PA-FGRS), validate it under simulations, and apply it to study MDD in the iPSYCH2015 case-cohort study. We demonstrate that combining PA-FGRS with genotypes improves three lines of inquiry: 1) classification of MDD in the context of PGS, 2) identifying robust genetic contributions to clinical heterogeneity of MDD, and 3) improving power in large single cohort GWAS of MDD. Our applications confirm, add context to, and extend recent methodological advances and their applications in similar data. The PA-FGRS framework is extensible, powerful and well-calibrated and could be applied to large biobanks or smaller family studies to pursue similar aims with other complex disorders.

## Results

### The iPSYCH 2015 MDD case-cohort genealogies are complex and contain a wealth of information

The iPSYCH 2015 case-cohort study ascertained 141,265 probands from the population born in Denmark between May 1st 1981 and Dec 31st 2008 (N=1,657,449) by cross-linking the Danish Civil Registration System^22^ (CPR) and The Danish Neonatal Screening Biobank^23^. The CPR includes all individuals who have legally resided in Denmark since its establishment in 1968 and each proband is associated with parental identifiers, where known. We use mother-father-proband connections^24^ to reconstruct extended genealogies (Online Methods) of 141,265 iPSYCH2015 probands, identifying 2,066,657 unique relatives spanning up to nine generations (birth years range: 1870s to 2016; Figure 1A, Supplementary Figure S1). Of 20,071,410 relative pairs identified, 24,773 pairs included two iPSYCH probands genotyped on the same array. Pedigree-inferred kinship was highly correlated with SNP-based kinship (r=0.969, Figure 1B), siblings sharing one recorded parent (with the other missing) tended to be half-siblings (Supplementary Figure S2), and approximately 45% of same-sex twins were monozygotic (Supplementary Figure S3). For genealogies of 141,265 probands included 99.5% of parents, 82.0% of grandparents, and 7% of great-grandparents, with the number of relatives identified per proband varying considerably (Figure 1C). Clinical diagnoses are aggregated for all relatives during periods of legal residence within Denmark from 1968 with in-patient psychiatric contacts recorded from 1969 to 1994 using ICD-8 and ICD-10 from 1994 onwards, and since 1995 both in- and out-patient contacts recorded (Figure 1D,E). There is a wealth of high-quality psychiatric familial phenotypes for each genotyped proband (Figure 1), but relatives are neither completely nor consistently observed.

**Figure 1.**
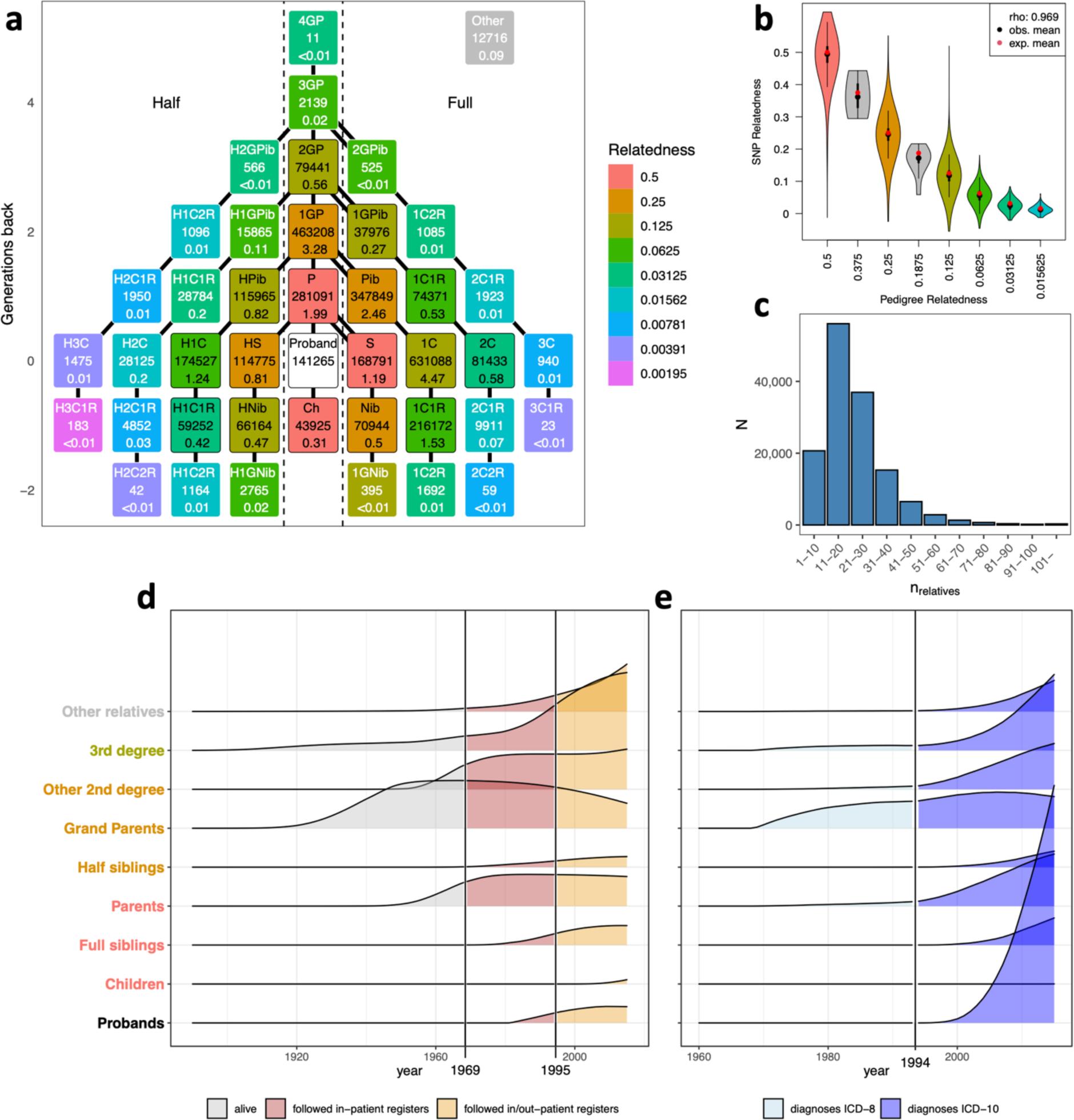
The iPSYCH 2015 MDD case-cohort genealogies are complex and contain a wealth of information. **a**) Each of the 141,265 probands (white box) in iPSYCH2015 can be connected to a number of different types of relatives, here reported as a total across all probands and average per proband. **b**) SNP-based relatedness is highly correlated with that inferred from genealogies. **c)** The number of relatives linked to each proband varies considerably. **d**) The proportion of total person-years of follow up is distributed differently across probands and their relatives, showing variability by relative type (y-axis), year of observation (x-axis), and register era (color). **e**) The proportion of total person-years of follow up for MDD cases similarly varies. P, parents; S, siblings; Ch, children; 1GP, grandparents; Pib(lings), aunts and uncles; Nib(lings), nieces and nephews; iCjR i^th^ cousin, j^th^ removed; H-, half; Other, relative types not in the figure. MDD, Major Depressive Disorder.

### PA-FGRS is a flexible, powerful framework for estimating individual liability scores

PA-FGRS estimates the expected genetic liability carried by a proband from an arbitrary set of relatives, assuming the outcome results from a thresholded latent Gaussian liability (Figure 2). As input PA-FGRS takes a kinship matrix, diagnostic status and age (at censoring, diagnosis, or end of follow-up) for each relative, disorder heritability, and individual morbid risks which may be estimated from lifetime sex by birth year-specific cumulative incidence. In a first step, each pedigree member is assigned an initial liability of 0 with variance 1. Then we consecutively condition on observations of other relatives, 1, …, n, updating all expected liabilities based on each relative. We first update the expected liability of a selected relative, r_(i)_, estimating their expected liability given their prior liability distribution, disease status, age and the lifetime incidence estimate. Then we update the liabilities of all remaining relatives, r_i+1_, …, r_n_, according to the PA-selection formula^19^ and a modified kinship matrix (Supplementary Figure S4). An optional final step updates the proband liability on their own diagnostic status and age. PA-FGRS produces a continuous score that summarizes the genetic liability from the proband’s pedigree.

**Figure 2.**
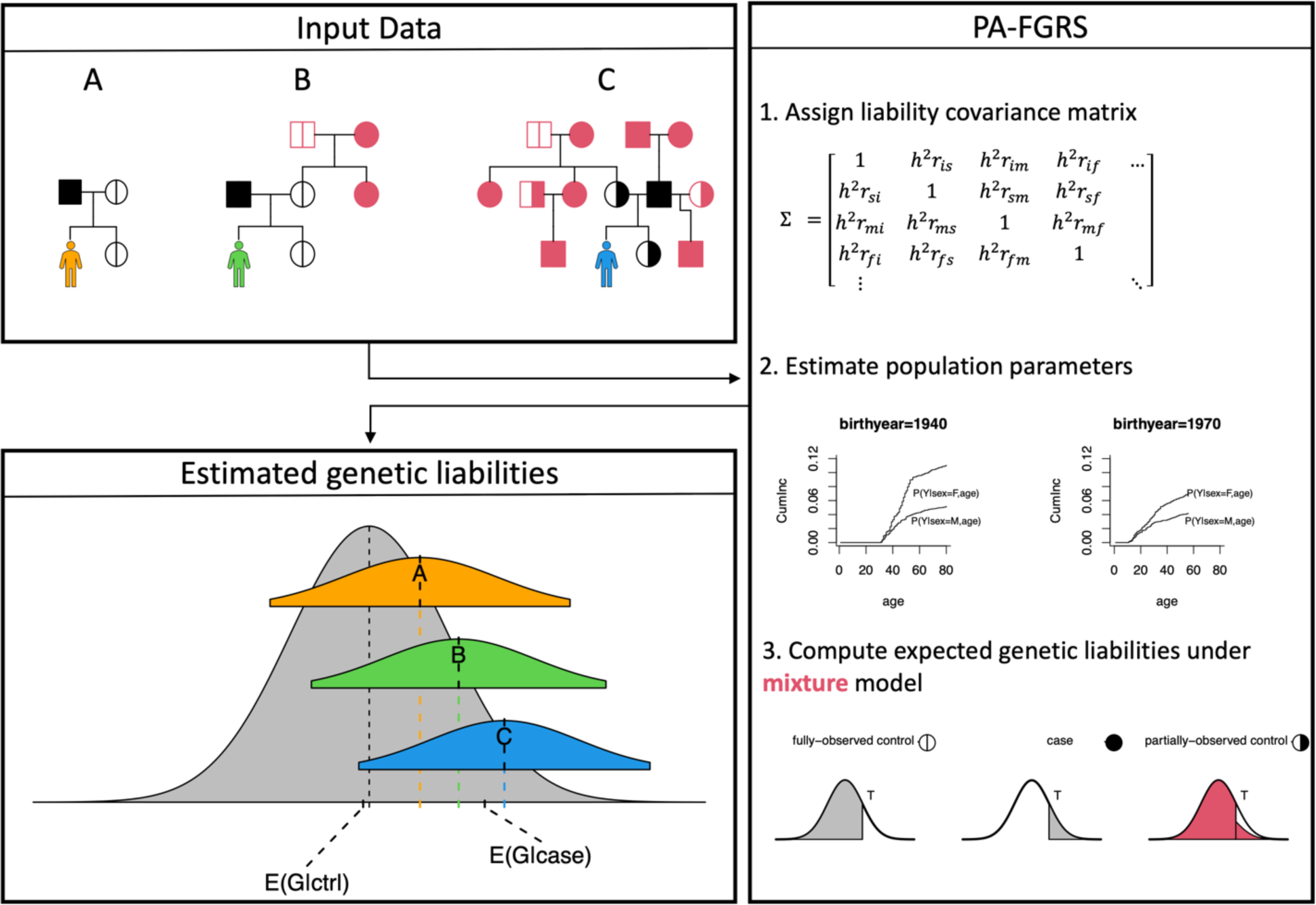
PA-FGRS estimates a continuous liability score for a proband from diagnoses in relatives and specific population parameters. PA-FGRS estimates latent disease liability in a proband from patterns of diagnoses in arbitrarily structured pedigrees where relative phenotypes may be age-censored. *Input data* for a proband can be a simple, fully-observed pedigree (i.e., no censoring; yellow proband), an extended pedigree with fully observed phenotypes (green proband), or an arbitrarily structured pedigree where many relative phenotypes may be age-censored (blue proband). *PA-FGRS* combines (1) an assumed form for covariance in liabilities among relatives with (2) estimates of individual morbid risk from, e.g., covariate stratified cumulative incidence curves, in (3) a novel extension of the Pearson-Aitken selection formulas that models age-censored controls as a mixture of cases and controls. *Estimated genetic liabilities* are assigned to each proband and determined by the unique configuration of their pedigree, population parameters, and the morbid risk of each relative. Proband liabilities (colored) are shown against the population distribution of genetic liability (gray) with E(G|case) and E(G|ctrl) indicating the expected population (i.e., unconditioned) mean liability of a case and control, respectively.

Other methods have approached this problem, but with limitations critical to our intended use case. Prior implementations^18, 25^ of the Pearson-Aitken (PA) selection formula^19^ assumed no age-censoring, which we address by modeling individuals as a mixture of truncated Gaussians, with mixture proportions reflecting individual morbid risks (Online Methods). FGRS^21^ followed this concept, but PA-FGRS takes a more formal approach that incorporates kinship relationships among relatives as well as between relatives and proband, producing better calibrated scores and estimates of conditional liability variance (Online Methods).

### Simulations demonstrate the advantages of PA-FGRS over other methods

We simulated 6,750,000 four-generational pedigrees with an average of nine relatives per proband (range 0-18), generating phenotypes from a liability threshold model (Online Methods). We found five considered methods, PA-FGRS, FGRS^21^, PA^18^, LT-PA^7^, and a fully specified Gibbs sampling-based approach^20^, gave estimates that were highly correlated (Figure 3A,B, *r* > 0.8), suggesting that they target similar latent constructs. Methods incorporating more similar information were more highly concordant, e.g., extended relatives (Figure 3A,B, *r* > 0.89) or extended relatives and censoring (Figure 3A, *r* > 0.95). The Gibbs sampling approach^20^ produced nearly identical estimates to PA-FGRS (*r*=0.999, Figure 3A,B), suggesting PA-FGRS behaves near optimally. Although similar, PA-FGRS consistently produced the highest correlations with true liability across a range of simulated heritabilities and prevalences (Figure 3C,D). The largest relative gains were when heritability, prevalence, and censoring were high. PA-FGRS was also the only method that was well-calibrated in the presence of censored data (Figure 3E; without censoring: Supplementary Figure 5). Our implementation of the Gibbs-sampling approach is computationally intractable at scale, limiting applicability. (Supplementary Figure 6)

**Figure 3.**
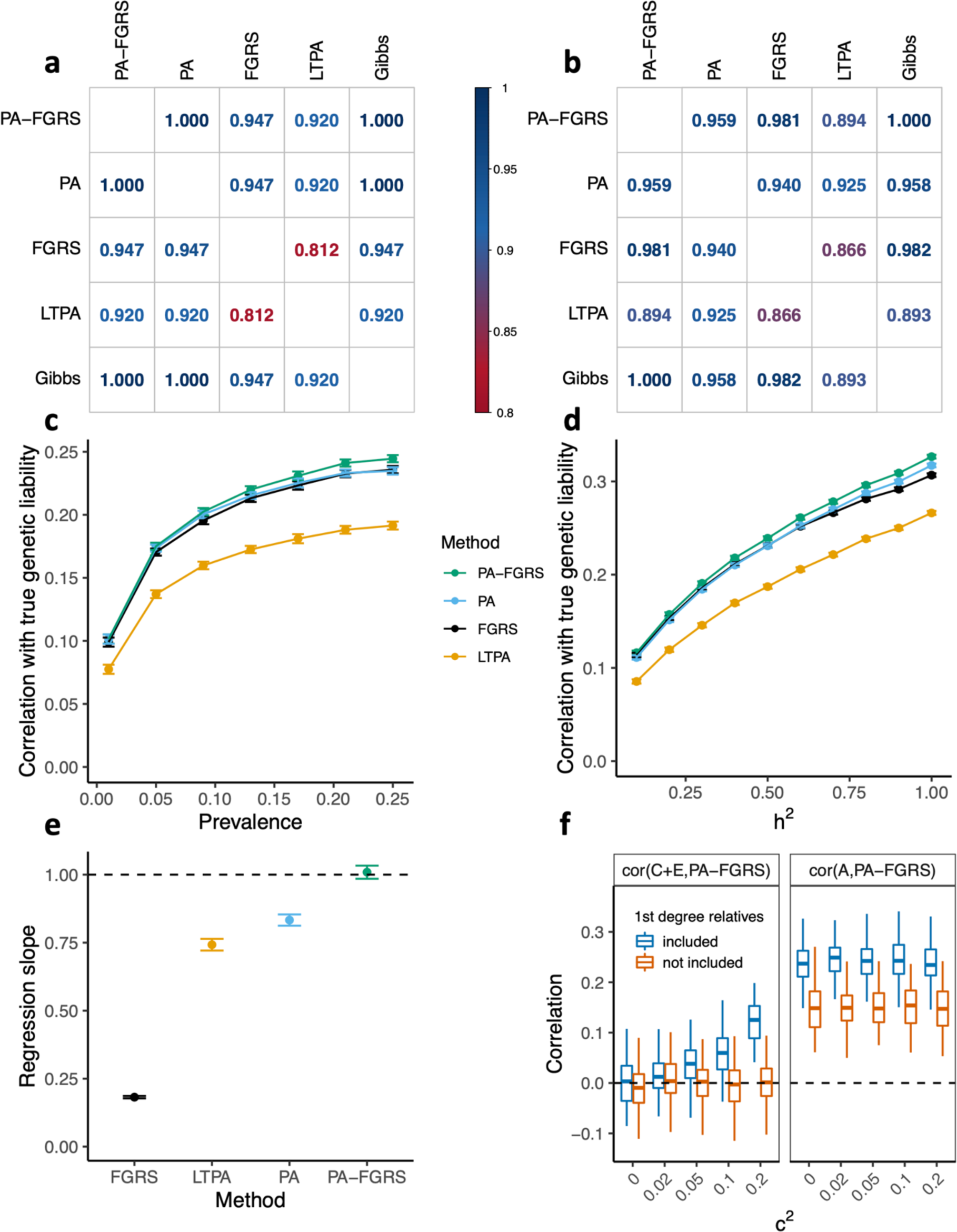
Simulations demonstrate the advantages of PA-FGRS over other methods. PA-FGRS liabilities are correlated with those from other methods in simulations (**a**) when all relatives are fully observed or **b**) when younger relatives are age-censored. In simulations, PA-FGRS shows the largest correlation with true genetic liability in simulations where age-censoring is present across **c**) varying trait prevalence and **d**) varying trait heritability. **e**) Linear regression of estimated liability on true liability shows PA-FGRS estimates are, uniquely, calibrated in the presence of age-censored records. **f**) The presence of shared environmental effects in generative models of familial resemblance creates correlation between PA-FGRS and this environmental component of liability. This can be reduced at the cost of power (i.e. reduced correlation with genetic components) by excluding confounded (i.e., first-degree) relatives. Panels c-e show mean and 95%-confidence interval across simulations, while f shows median, range and interquartile range across simulations.

One limitation of methods that consider only first-degree relatives^7, 17^ is that estimated genetic liabilities may be unduly influenced by effects of familial environment. This may be desirable if the goal is to optimize prediction^9, 18^, only, but less so if the goal is to make etiological inferences^21^. We repeated our simulations including a common environment component of variance shared among first degree relatives (Figure 3F, Supplementary Figure 7) - a typical quantitative genetics model^26^. PA-FGRS (and all other approaches) produce liability estimates that are correlated with environmental liability (Figure 3F). With extended genealogies we can omit close relatives as a sensitivity test for undue influence. Liabilities estimated after excluding first degree relatives remained good estimators of genetic liability and were uncorrelated with environmental liability (Figure 3F). The flexibility of PA-FGRS can add important context to estimated liabilities that may be especially important when interpreting, e.g., profiles of liability scores^21, 27^ or if shared environment is a concern.

PA-FGRS requires external estimates of specific population parameters, namely, lifetime prevalence and heritability. Providing inaccurate estimates leads to miscalibrated liabilities, but has modest impact on the correlation between estimated and true liability in simulations (Supplementary Figure 8).

### PA-FGRS contribute to classification models of MDD over and above PGS

Both family history and PGS are explain liability for MDD. Using a two-fold split of iPSYCH (Supplementary Figure 9), we trained a model to classify MDD from combinations of PA-FGRS and PGS in iPSYCH 2012 (or iPSYCH 2015i) and evaluated classification accuracy in the complement, iPSYCH 2015i (or iPSYCH 2012; Online Methods, Figure 4A,B, Supplementary Table S6). Both genetic instruments, fit alone, significantly classify MDD cases from controls, in both cohorts: iPSYCH2012 (AUC_PGS_=0.588 (0.583-0.594), p= 3.7 × 10^−229^; AUC_PA-FGRS_=0.598 (0.592-0.603), p=4.9 × 10^−328^) and iPSYCH2015i (AUC_PGS_=0.573 (0.565-0.580), p=7.8 × 10^−94^; AUC_PA-FGRS_=0.576 (0.569-0.583), p=4.1 × 10^−136^). When combined in a multivariate model, each genetic instrument contributes independent information to classification, with combined effects of PA-FGRS and PGS larger than individual effects (iPSYCH2012: AUC_PGS+FGRS_=0.630 (0.625-0.638) and iPSYCH2015i: AUC_PGS+FGRS_=0.608 (0.601-0.615)).

**Figure 4.**
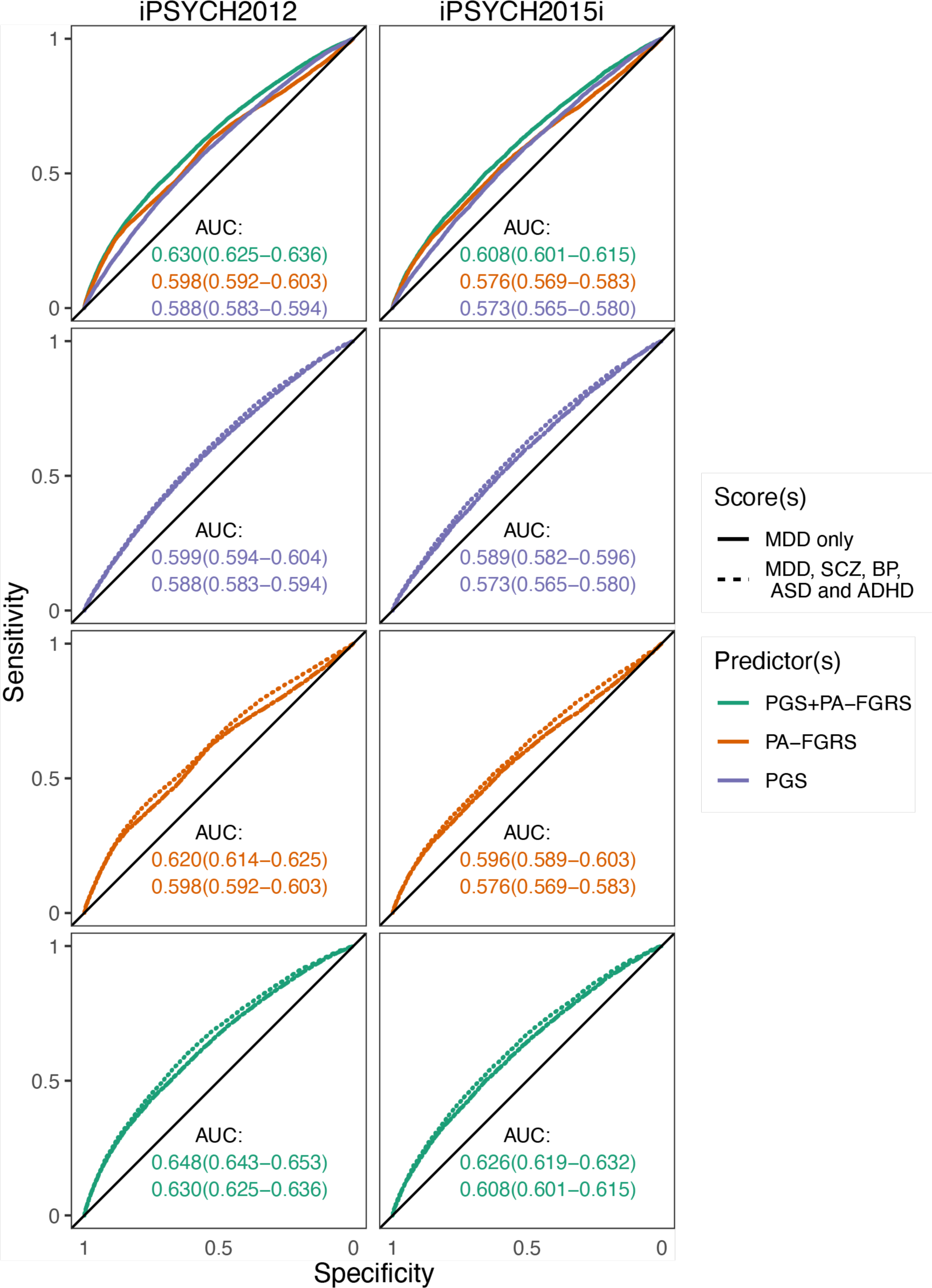
PA-FGRS contribute to classification models of MDD over and above PGS. Combining PA-PGRS-MDD and PGS-MDD improves classification of MDD (**a)** the iPSYCH-2012 (Ncases=20,632, Nctrl=23,870) and (**b)** iPSYCH-2015i (Ncases=10,317, Nctrl=15,785) case-cohorts. Using PGSs for five disorders improves prediction of MDD over only PGS-MDD in **(c)** iPSYCH-2012 and **(d)** iPSYCH-2015i. Using PA-FGRS for five disorders improves prediction of MDD over only PA-FGRS-MDD in **(e)** iPSYCH-2012 and **(f)** iPSYCH-2015i. Combining PA-FGRS for five disorders with PGSs for five disorders improves prediction of MDD over only PA-FGRS-MDD and PGS-MDD in (**g**) iPSYCH-2012 and (**h**) iPSYCH-2015i . **AUC** area under the receiver operating characteristic curve with 95%-confidence interval.

Including PGS for four other psychiatric disorders, schizophrenia (SCZ), bipolar disorder (BPD), autism spectrum disorder (ASD), and attention deficit/hyperactivity disorder (ADHD), improved the classification of MDD relative to models with MDD PGS only (iPSYCH2012: AUC_5-PGS_=0.599 (0.594-0.604); iPSYCH2015i: AUC_5-PGS_=0.589 (0.582-0.596); Figure 4C,D, Supplementary Table S6). Similarly, incorporating PA-FGRS for the four other psychiatric disorders improved the classification of MDD relative to models with MDD PA-FGRS only (iPSYCH2012: AUC_5-PA-FGRS_=0.620 (0.614-0.625); iPSYCH2015i: AUC_5-PA-FGRS_=0.596 (0.589-0.603), Figure 4E,F). Combining all 10 predictors resulted in the best out of sample classification (iPSYCH2012: AUC_5-PGS+5-PA-FGRS_=0.648 (0.643-0.653); iPSYCH2015i: AUC_5-PGS+5-PA-FGRS_=0.626 (0.619-0.632), Figure 4G,H). These results demonstrate that combining genetic instruments that leverage different sources of genetic information improves classification of MDD.

### Composite genetic profiles identify robust signatures of genetic heterogeneity in MDD

Individuals diagnosed with MDD demonstrate extensive clinical heterogeneity that may reflect etiologic heterogeneity. We used multinomial logistic regression to associate differences in clinical presentations of individuals diagnosed with MDD to genetic liability profiles (Online Methods, Figure 5). We leverage the complementarity of PGS and PA-FGRS, above, by defining composite genetic liability scores (e.g. BPD-score= *β*_PGS_*PGS_BPD_+ *β*_PA-FGRS_*PA-FGRS_BPD_, where *β*_PGS_ and *β*_PA-FGRS_ are the estimated effect of the PGS and PA-FGRS on their natural outcome in a case-control logistic regression). Each composite liability score was significantly larger in individuals diagnosed with MDD than in controls, across all subgroups (Figure 5; p<0.05). The liability scores for bipolar disorder (BPD), schizophrenia (SCZ), autism spectrum disorders (ASD) and attention deficit/hyperactivity disorder (ADHD) tended to have smaller effects on MDD subgroups than on their natural outcome (i.e., β_MLR_/β_LR_ < 1; the colored bars below dashed line in Figure 5; Online Methods), except for BPD liability on conversion to a BPD diagnosis (β_MLR_/β_LR_ =0.97 (0.90-1.04), Figure 5A).

**Figure 5.**
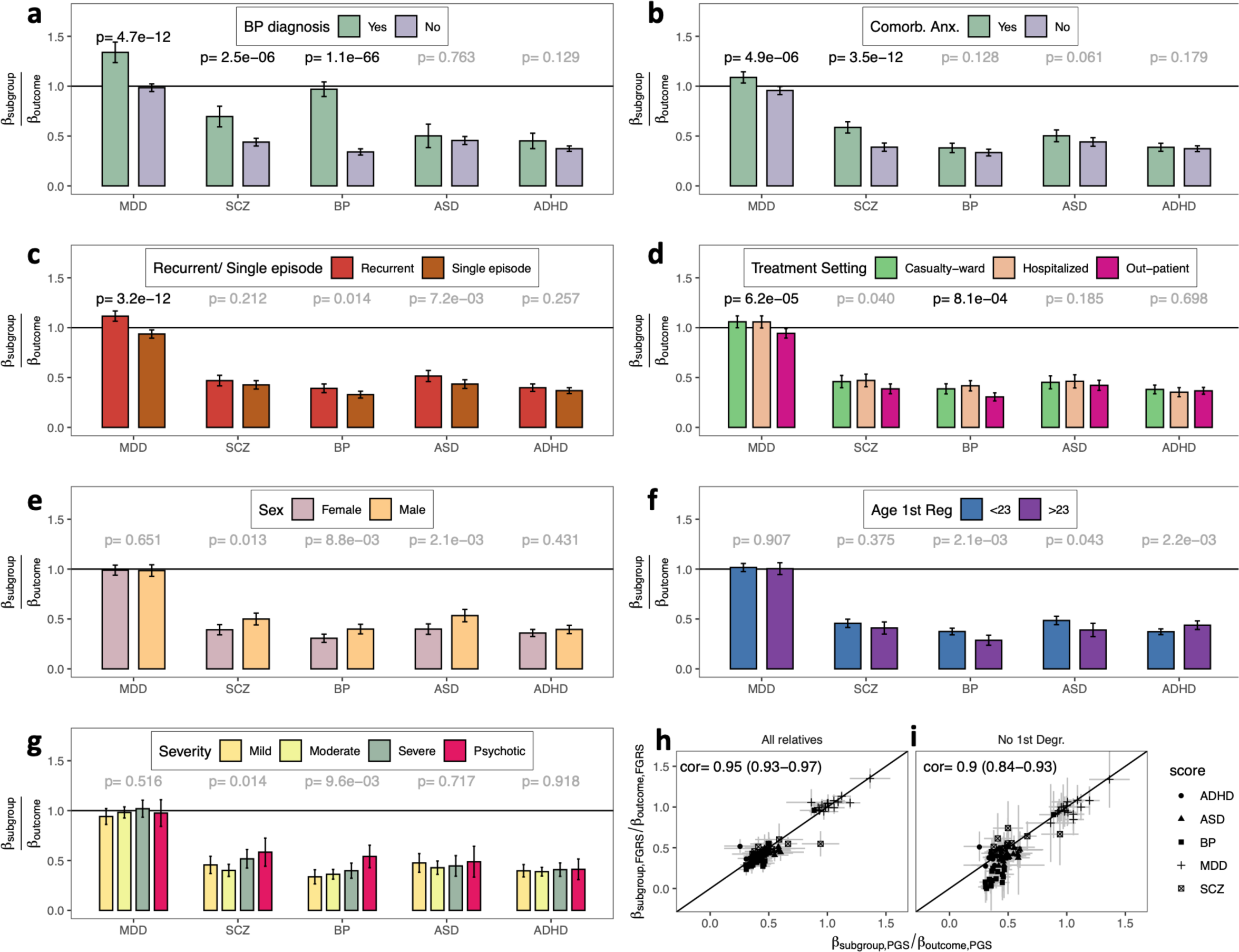
Composite genetic profiles identify robust signatures of genetic heterogeneity in MDD. We predicted MDD subgroup membership from composite genetic liability scores that integrates PGS and PA-FGRS, together, in multinomial logistic regression with controls as a reference group. (**a**) Higher MDD, SCZ, and BPD genetic liability were associated with conversion from MDD to BPD. (**b**) Higher MDD and SCZ genetic liability were associated with a comorbid anxiety diagnosis. (**c**) Higher MDD genetic liability was associated with recurrent MDD (**d**) lower MDD and BPD genetic liability were associated with out-patient treatment. No differences were observed (**e**) between females and males diagnosed with MDD, (**f**) first MDD diagnosis before/after age 23, or (**g**) mild, moderate, severe, or psychotic depression. PGS-only and PA-FGRS-only effects are highly consistent both, when **(h)** using all relatives and **(i)** when excluding first degree relatives. **(a-g)** Effect sizes are presented on a calibrated scale, where the regression coefficient describing the effect of a genetic liability score on the subgroup is divided by the coefficient of the same score when predicting its natural outcome (i.e., BPD score predicting BPD) in a simple logistic regression. This places the magnitude of subgroup effects on a scale that is relative to the effect of the score in its distinguishing natural outcome from controls, which can account for differences in the sensitivity of the individual scores. **(a-f)** models are meta-analyzed across iPSYCH2012 (Ncases≤20,632, Nctrl≤23,870) and iPSYCH2015i (Ncases≤10,317, Nctrl≤15,785), **(g)** is only available in iPSYCH2012. Significance is depicted in bold, at p < 0.05 / 35. Detailed sample sizes for the individual analyses are provided in Table S3. Error-bars indicate 95% confidence intervals. MDD major depressive disorder; SCZ schizophrenia; BPD bipolar disorder; ASD autism spectrum disorders; ADHD attention-deficit/ hyperactivity disorder; p, p-value.

Among 30,949 individuals diagnosed with MDD, those also diagnosed with BPD (N=1,477) had significantly (*p* < 1.4 × 10^-3^, adjusting for 35 tests) higher genetic liability for MDD (*p* = 1.1 × 10^−12^), BPD (*p* = 4.7 × 10^−66^), and SCZ (*p* = 2.5 × 10^−6^; Figure 5A). Among the 29,472 individuals diagnosed with MDD (excluding BPD) the 7,205 also diagnosed with an anxiety disorder had higher genetic liability to MDD (*p* = 4.9 × 10^−6^) and SCZ (*p* = 3.5 × 10^−12^; Figure 5B). Individuals with recurrent depression (N=9,903) had higher liability to MDD (*p* = 3.2 × 10^−12^; Figure 5C) than those with single episode depression (N=19,569). Individuals treated for MDD in-patient (N_Hospitalized_=5,815) had higher liability to MDD (*p* = 6.2 × 10^−5^) and BPD (*p* = 8.1 × 10^−4^) than those treated out-patient (N_Out-patient_=12,432, Figure 5D). We did not observe any significant differences (*p* > 1.4 × 10^−3^) in the genetic liability score profiles of males vs females (N_Female_=19,906, N_Male_=9566; Figure 5E), based on age-at-first-diagnosis (Figure 5F), or based on diagnostic codes for severity (mild N_Mild_=3,004, N_Moderate_ =8,742, N_Severe_= 2,391, N_Psychotic_=856; Figure 5G).

Each analysis was repeated using PGS- or PA-FGRS-only profiles (Supplementary Figures S10-S11). PGS-only and PA-FGRS-only results were highly similar (r=0.95 (0.93-0.97); Figure 5H) and PA-FGRS or PGS scores, alone, were less powerful than composite scores (PA-FGRS-only mean log_10_(p)= 2.90; PGS-only mean log_10_(p)= 2.47; composite mean log_10_(p)= 4.24). PGS and PA-FGRS appear to capture similar constructs and by combining the two we can increase power to detect genetic heterogeneity. Finally, to test for large effects of the familial environment, we constructed PA-FGRS excluding nuclear family members (i.e., parents, siblings, half-siblings, and children). The overall trends were highly consistent with the full analysis (Figure 5I), albeit with reduced significance (Supplementary Figure 12). Genetic liability score profiles are associated with differences in the clinical presentation of MDD, involving contributions from non-MDD liability scores, with parallel trends in PGS or PA-FGRS alone, and do not seem strongly influenced by familial environment.

### GWAS on PA-FGRS liability values adds power to single cohort MDD GWAS

Studying genetic liability of threshold trait is expected to boost power in GWAS (Supplementary Figure 13). We performed meta-analytic GWAS across the iPSYCH 2012 (N_cases_=17,518, N_ctrl_=23,341) and 2015i (N_cases_=8,323, N_ctrl_=15,204, Supplementary Figure 9) cohorts and compare logistic regression GWAS of binary diagnoses to linear regression GWAS of PA-FGRS (Online Methods, Figure 6). GWAS of PA-FGRS identified 3 independent loci (Figure 6A; index SNPs: rs16827974, *β*=0.014, p=2.9 × 10^−8^; rs1040574, *β*=-0.011. p=3.3 × 10^−8^; rs112585366, *β*=0.026, p=4.4 × 10^−8^; Supplementary Table S7). These three variants and 24 of the 29 suggestive loci (false discovery rate <0.05) showed consistent sign in an independent MDD GWAS from Howard el al.^28^ (excluding iPSYCH, Supplementary Table S2-S3). GWAS of binary diagnoses identified two of these loci (Figure 6B; index SNPs: rs6780942, 8.5Kb from rs16827974 Beta=0.085, p=7.1 × 10^−9^; rs3777421 36.3Kb from rs1040574, *β*=-0.073, p=4.6 × 10^−8^, Supplementary Table S8). These two and 24 of the 35 suggestive loci (false discovery rate <0.05) showed consistent sign in Howard el al.^28^ (excluding iPSYCH, Supplementary Table S2-S3). The 28 independent, genome-wide significant index SNPs reported in Howard el al.^28^ (excluding iPSYCH) have slightly, but significantly, larger test statistics in the GWAS on PA-FGRS (PA-FGRS mean χ^2^= 4.55; case-control mean χ^2^= 3.80; paired t-test p=0.018; Figure 6C).

**Figure 6.**
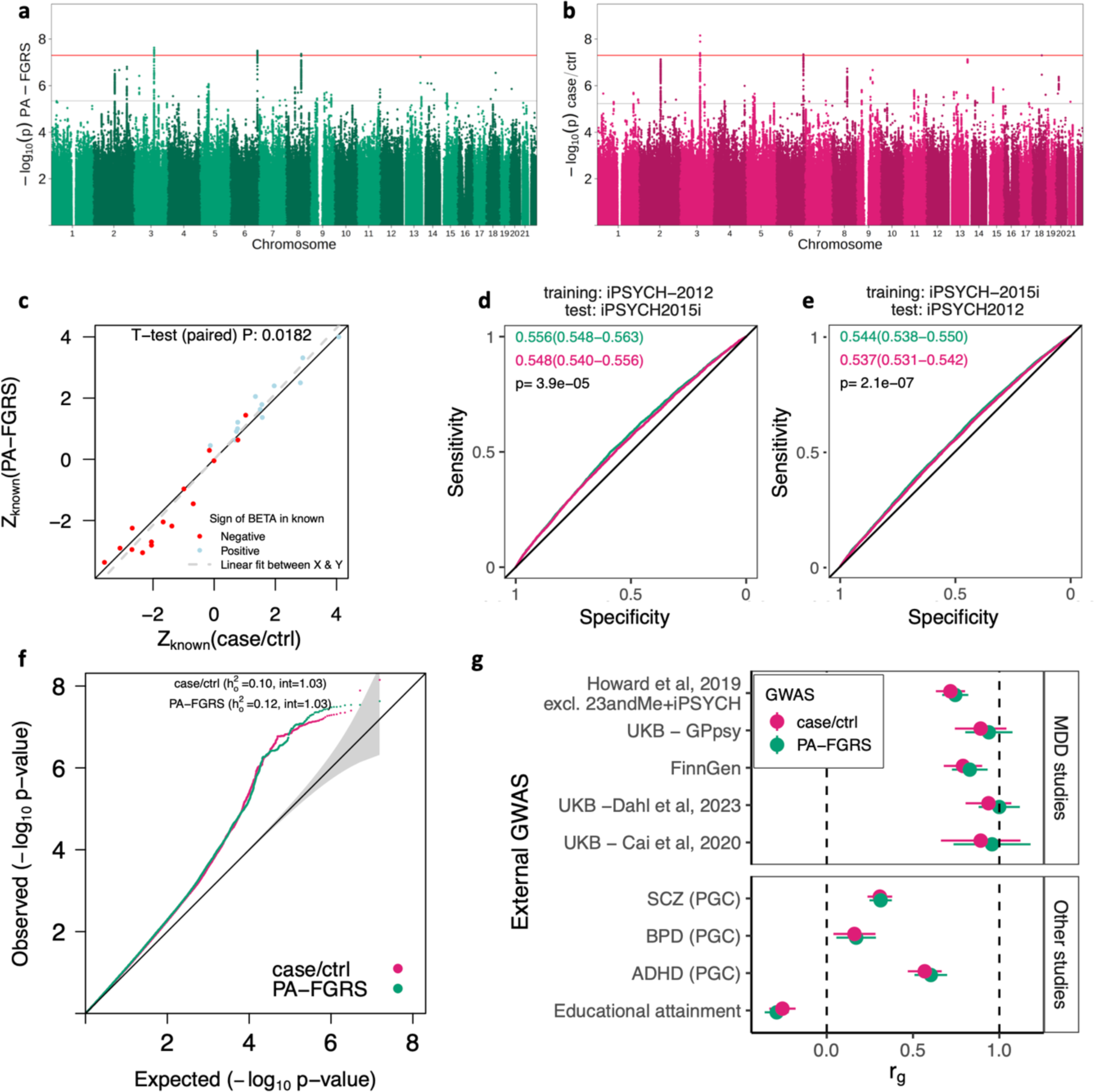
PA-FGRS liabilities improve power for GWAS of MDD. Genome wide association studies (GWAS) of 25,841 cases and 38,545 controls using (**a**) PA-FGRS liability finds three independent genome-wide significant loci, while (**b**) logistic regression (case/control) finds two. **c)** PA-FGRS GWAS test statistics are more extreme (i.e., more significant) than case-control GWAS at index SNPs of 28 loci reported in a previous GWAS of MDD. PGS trained using PA-FGRS GWAS achieve higher classification accuracy that those trained on case-control GWAS in **d)** iPSYCH2012 and **e)** iPSYCH 2015i, two independent evaluation cohorts. **f)** SNP-heritability estimated by LD-score regression analyses is slightly, but not significantly, larger for PA-FGRS GWAS, while estimated intercepts are equivalent. **g)** PA-FGRS and case-control GWAS show similar genetic correlations with external GWAS of MDD and related traits.

Next, we trained polygenic scores in each subcohort (iPSYCH2012 or iPSYCH2015i) using GWAS performed in the other (iPSYCH2015i or iPSYCH2012). In both cohorts, PGS trained with PA-FGRS GWAS were modestly, but significantly, better at classifying MDD versus controls (2012: AUC_case-control PGS_ =0.537 (0.531-0.542), AUC_PA-FGRS PGS_ =0.544 (0.538-0.550), test of differences: p=3.9 × 10^−5^; 2015i: AUC_case-control PGS_ =0.556 (0.548-0.563), AUC_PA-FGRS,PGS_=0.548 (0.540-0.556), test of differences: p=2.1 × 10^−7^; Figure 6D). Observed scale SNP-h^2^ was larger in the PA-FGRS GWAS, but this difference was not significant (h^2^_obs,PA-FGRS_-h^2^_obs,PA-case/ctrl_=0.015 (-0.013-0.043); Figure 6E), and genetic correlations with external studies of MDD and other psychiatric disorders were similar (Figure 6F).

## Discussion

We have developed a new method for estimating genetic liability, PA-FGRS, that is more generalizable across data sets and research questions and outperforming existing methods in complex genealogical data. We show that PA-FGRS complements genotype-based inferences into MDD in three ways: 1) PA-FGRS liabilities improve classification models when fit together with state-of-the-field PGS, 2) combing PA-FGRS and PGS better describes the etiology underlying clinical heterogeneity associated with comorbidity, recurrence, and severity in MDD, 3) GWAS performed on PA-FGRS scores have more power than GWAS on case-control status. Our method is flexible, easy to use, and could be applied to ask similar questions of other complex diseases. Our data-first approach - describing the unique characteristics of a powerful resource and tailoring a novel method to accommodate its peculiarities - allows us to leverage, rather than discard or censor inconvenient data. This is a complementary approach to lowest common denominator cross-cohort studies, and may be especially relevant as newer, larger, deeper, and necessarily more peculiar, data emerge.

PA-FGRS is unique in that it is model based, incorporates distant relatives, *and* handles age-censored phenotypes of relatives. Incorporating distant relatives allows us to manipulate our liability calculation to exclude close relatives as a sensitivity test for undue impact of familial environment. This, and using morbid risk to define a mixture model, makes PA-FGRS most similar in concept to FGRS^21^, but the formal model underlying PA-FGRS (PA-selection theory) gains efficiency, and improves calibration of estimated liabilities. The way we model censoring also makes the underlying etiological model assumed by PA-FGRS qualitatively different from, e.g., LT-FH++^17^ or ADuLT^29^. PA-FGRS assigns the same liability to rare cases as random cases (i.e., uses one population threshold), using covariate stratified cumulative incidence to define mixture proportions for controls. LT-FH++ and ADuLT assign increasingly larger genetic liability to increasingly rarer cases observed in empircal cumulative incidence curves (i.e., uses per individual thresholds). For example, in our MDD data, males diagnosed at a young age, many decades back (a rare event in empirical cumulative incidence) would make much larger contributions to liability estimates in the LT-FH++/ADuLT model than in the PA-FGRS framework. The underlying model of PA-FGRS is amenable to analysis and extension, representing an advantage for future work that could extend the model to include non-additive covariance, multiple traits, or more complicated etiological models of heterogeneity and comorbidity.

Combining family based liabilities and genotype-based PGS from multiple disorders significantly improved classification accuracy. In cancer^30^ or coronary artery disease^31^, risk models incorporate multiple measures - health states, health traits, family history, and PGS. In psychiatry, this has been pursued in more limited contexts, (e.g.,^8^). Previous studies have found that combining parental history information and PGS improves the prediction accuracy^8^, however, these studies only considered risk associated with parental MDD and did not leverage diagnoses in other relatives. Integrative models that combine multiple sources of genetic information, such as family history, estimated liability, and PGS along with exposure data have the potential to advance the clinical utility of risk assessment in psychiatry but will require large population data and integrative models.

Our novel composite profile analysis replicates, extends, and adds context to previous work considering genetic heterogeneity in MDD. First, we replicate previous results in similar data showing statistically significant associations of genetic liability to MDD with recurrence and MDD and BPD with treatment location^3233^. Our models calibrate effect sizes differently to accommodate noisy instruments, such as PGS, leading to different framing of effect sizes, which may be moderate to substantial, rather than minimal. We also replicate associations between BPD and SCZ liability and conversion from MDD to BPD^34^, but interpret BPD genetic liability to be significantly more important than SCZ genetic liability for conversion. Second, studies in Swedish registers have shown differences in family genetic risk scores (FGRS) associated with progression to BPD^21^, comorbid anxiety^27^, recurrence^21^, treatment setting^35^, and age-at-onset^21^ of MDD. We confirm higher genetic liability to MDD among cases with recurrent depression using composite, PGSs-only, and PA-FGRS-only liability scores. We also replicate a higher liability to MDD and SCZ among MDD cases with comorbid anxiety using our composite and PGS-only scores, and saw the same trend of higher liability to MDD and BPD among hospitalized cases using our composite and PGS-only scores. Findings of higher BPD liability in male MDD cases^35^ was nominally significant in our study. We did not observe associations to age at onset, however, iPSYCH has a reduced range for onset (15 to 35 vs. <22 to >69) and includes secondary care treated (i.e., more severe) MDD. Our study replicates and extends previous results by providing more interpretable effect sizes, using an alternative model-based approach for family liability scores, and by showing consistency between familial and molecular scores.

We observed a small, significant improvement in power when performing GWAS for MDD on PA-FGRS liabilities. A previous study incorporating family history in GWAS of MDD using iPSYCH did not observe gains^17^, but only considered first degree relatives and weighted them differently. Consistent with simulations, the relative increase in power observed in highly ascertained case-control data is smaller than what has been reported for population-based studies^7, 17^. In population studies, especially for rarer disorders, most of the variance in liability is hidden *within controls*, whereas for highly ascertained data, most of the variance in liability remains *between cases and controls*. In this latter context, little is gained by moving from binary to continuous measures. Although we observe small gains in power for GWAS, consistent with other studies, the most impactful applications of PA-FGRS may lie in classification and descriptions of etiology.

Our study should be interpreted in light of a few important limitations. Certain modeling choices could affect the reliability of PA-FGRS. First, pedigree size varies substantially among individuals. Probands with few relatives have scores regressed more towards the mean liability which can introduce bias. Second, modeling the censoring process requires external information about age-of-onset curves for disease of interest - as do the other methods - and these may change in calendar time cohorts. While reliable age-of-onset curves are available for the present register coverage, estimating age of onset curves for past decades, with different diagnostic systems and different register coverage is challenging. Third, our model assumes that the true liability of cases with different age and calendar year of onset is the same, while others have proposed true liability should vary according to these covariates^17, 29^. Both approaches are based on heuristics and could be better compared, integrated, and optimized to improve performance.

Here, we have taken a data-first approach to studying the genetic architecture of MDD by tailoring both our study aims and method development to the particular strengths and challenges of a unique data resource. Doing so resulted in a methodological increment with broad applicability and highlights the utility of integrating multiple sources of genetic data when considering trait predictions, etiological descriptions, and gene mapping.

## Online Methods

### iPSYCH 2015 case-cohort study

The Lundbeck Foundation initiative for Integrative Psychiatric Research (iPSYCH)^3, 4^ is a case-cohort study of all singleton births between 1981 and 2008 to mothers legally residing in Denmark and who were alive and residing in Denmark on their first birthday (N=1,657,449). The iPSYCH 2015 case-cohort comprises two enrollments from this base population. The iPSYCH 2012 case-cohort enrolled 86,189 individuals (30,000 random population controls; 57,377 psychiatric cases)^3^. The iPSYCH 2015i case-cohort expanded enrollment by an additional 56,233 individuals (19,982 random population controls; 36,741 psychiatric cases)^3, 4^. DNA was extracted from dried blood spots stored in the Danish Neonatal Screening Biobank^23^ and genotyping was performed on the Infinium PsychChip v1.0 array (2012) or the Global Screening Array v2 (2015i). Psychiatric diagnoses were obtained from the Danish Psychiatric Central Research Register (PCR)^36^ and the Danish National Patient Register (DNPR)^37^. Diagnoses in these registers are made by licensed psychiatrists during in- or out-patient specialty care but diagnoses or treatments assigned in primary care are not included. Linkage across population registers, to parents where known, and to the neonatal biobank is possible via unique citizen identifiers of the Danish Civil Registration System^22^. The use of this data follows standards of the Danish Scientific Ethics Committee, the Danish Health Data Authority, the Danish Data Protection Agency, and the Danish Neonatal Screening Biobank Steering Committee. Data access was via secure portals in accordance with Danish data protection guidelines set by the Danish Data Protection Agency, the Danish Health Data Authority, and Statistics Denmark.

### Genotyping and quality control

Genotype phasing, imputation, and quality control were performed in parallel in the 2012 and 2015i cohorts according to custom, mirrored protocols. Briefly, phasing and imputation were conducted using BEAGLEv5.1^38, 39^, both steps including reference haplotypes from the Haplotype Reference Consortium v1.1 (HRC)^40^. Quality control was applied prior to and following imputation to correct for missing data across SNPs and individuals, SNPs showing deviations from Hardy-Weinberg equilibrium in cases or controls, abnormal heterozygosity of SNPs and samples, genotype-phenotype sex discordance, minor allele frequency (MAF), batch artifacts, and imputation quality. Kinship was detected within and across 2012 and 2015i cohorts using KING^41^, censoring to ensure no second degree or high relatives remained. Ancestry was examined using the smartpca module of EIGENSOFT^42^, and multivariate PCA outliers from the set of iPSYCH individuals with both parents and four grandparents born in Denmark were excluded. In total, 7,649,999 imputed allele dosages were retained for analysis.

### iPSYCH 2015 case-cohort genealogies

All recorded relatives of probands in this iPSYCH 2015 case-cohort were obtained from the Danish Civil Registry^22^ using mother-father-offspring linkages. From the 141,265^43^ probands, we identified 2,066,657 unique relatives, assembling all relationships into a population graph using the *kinship2*^44^ and *FamAgg*^43^ packages where edges denoted membership in a recorded trio. The relatedness coefficient for each pair was calculated as a weighted sum of unique ancestral paths through the population graph (i.e. not including the same individual, except for the common ancestor). Each path in the sum was weighted by (0.5)^(number of edges in the path)^45^. The Danish Civil Registry does not contain information on zygosity for same-sex twins, but following analysis of the SNP-kinship of children of same-sex twins (Supplementary Figure S3) we assigned same-sex twins a relatedness coefficient of 0.75. Similarly, guided by analysis of siblings with missing paternal records (Supplementary Figure S2), we assigned maternal siblings with missing paternal records a relatedness coefficient of 0.25. 24,773 pairs of relatives from the population genealogy included two probands genotyped on the same genotype array. We used Pearson’s correlation of the graph-inferred kinship and SNP-inferred kinship using KING^41^ as an estimate of concordance and quality of inferred relationships.

### Pearson-Aitken Family Genetic Risk Scores (PA-FGRS)

PA-FGRS estimates a liability for disease carried by a proband from the observed disease status in a pedigree and under the assumption of a liability threshold model for the disease^46^. The method first estimates an initial liability for each relative and then uses the Pearson-Aitken selection formula to sequentially update the expected liability in the proband conditional on each relative.^46, 47^

We begin by assuming a disease, *D_i_* = 1, arises when an individual, *i*, carries a latent liability, *L_i_*, that surpasses some threshold, *t*. Liability, *L_i_*, can arise from additive effects (β_j_) of genetic factors (X_ij_), or environmental deviations (e_i_) and genetic contributions follow classic polygenic theory.^46, 47^ We can write a generative model:

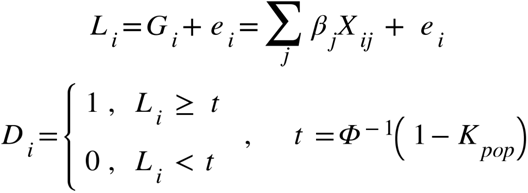

Where the threshold, *t*, is the standard normal quantile that corresponds to a cumulative probability of *k*_pop_, the lifetime prevalence of the disorder. Further we assume that the vector consisting of the genetic liability of the proband and the total liability of *n* genetic relatives [*L*_1_, . . ., *L_n_*, *G_p_*]^T^ ∼*MVN*([0, . . ., 0]^*T*^, Σ) with covariance matrix:

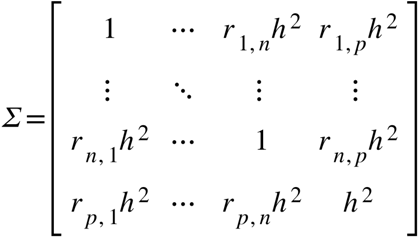

Under this model the expected value of *L*_i_, conditional on the true value for *D*_i_ is according to truncated normal distribution theory^48^:

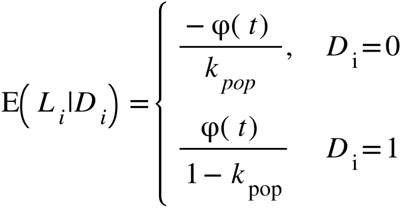

A critical assumption of this model is that each individual is fully observed (i.e., no age censoring), meaning there is an equivalence between their diagnostic and disorder status. This assumption rarely holds in practice, but the variable follow-up of relatives by the Danish register system makes it extremely tenuous. We instead propose a model (Supplementary Information) where the disease status *Y_i_* in those who surpass the threshold is observed with a probability corresponding to the ratio between (possibly stratified) age-specific prevalence (*K_i_*) and the life-time prevalence (*K_pop_*),

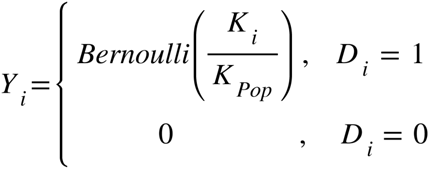

To get the expected liabilities under this model we use a mixture of an upper and a lower truncated Gaussian both with mean and variance corresponding to their conditional expectations, and with the mixture proportion (π_*i*_), corresponding to the conditional probability of being a case. Let *ψ*(μ, σ^2^, *a*, *b*) denote a truncated gaussian with mean μ, variance σ^2^, lower truncation at *a* and upper truncation at *b*. Then the distribution of *L_i_* conditional observations 1 to *i* is:

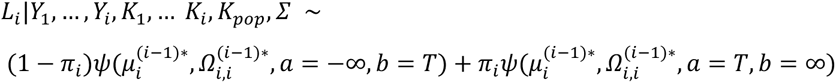

with 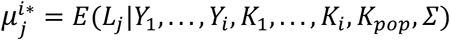 for *i*>0 and 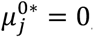, while 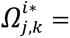 *Cov*(*L_j_*, *L_k_*|*Y*_1_,...,*Y_i_*, *K*_1_,...,*K_i_*, *K_pop_*, Σ) for *i*>0 and Ω^0*^ = Σ, and π_*i*_ = 1 if *Y_i_* = 1 and π_*i*_ P(*D_i_* = 1|*Y*_1_,..., *Y_i_*, *K*_1_,... *K_i_*, *K_pop_*, Σ) otherwise. This we approximate as (See Supplementary Information):

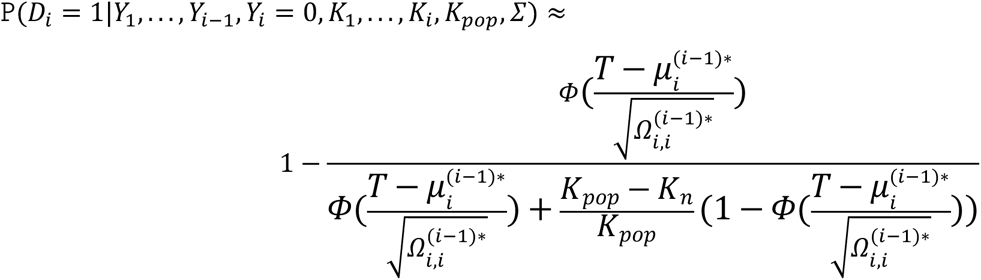

Following adaptations^18, 25^ of the Pearson-Aitken selection formula^49^ the conditional mean and variance of expected liability for a proband is estimated given their pedigree, initial liabilities, and population parameters^25^. 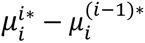 be the effect of conditioning on *Y_i_* and *K_i_* has on 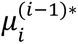 then the vector of conditional mean liabilities,μ^*i*∗^, is:

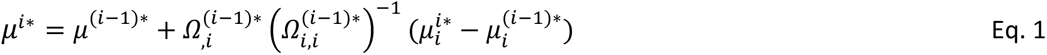

Similarly, if conditioning changes 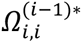 to 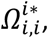, the conditional covariance matrix of liabilities, *Ω*^*i*∗^, is estimated as:

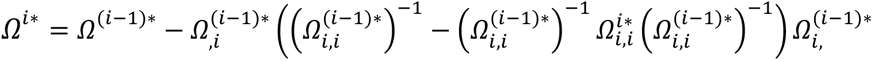

Previous work has found PA selection to be an efficient estimator of genetic liabilities of binary traits given family history.^7, 18, 25^ In practice, we start by setting the liability vector to a zero vector, we then iteratively condition on the observed disease status of each relative using the expected mean and a variance of a mixture of truncated gaussians in combination with the Pearson-Aitken selection formula to obtain the expected genetic liability of the index individual.

Our PA-FGRS is available as R code: https://github.com/BioPsyk/PAFGRS.

### Simulations

We simulated 500 proband pedigrees with varying numbers and kinds of relatives with variable age censoring. The heritability was set to 0.50 and prevalence to 0.4. We assessed the correlation between the estimated liabilities obtained from five different liability estimation methods: PA-FGRS, FGRS^21^, PA^18^, LT-PA^7^, and a Gibbs sampling-based approach^20^. Next, we repeated the simulations 4000 times with varying prevalence and 5000 times with varying heritability.

To assess the impact of shared environment (c^2^), we considered a generative model with an additional factor that determined the similarity between parents, off-spring and siblings (Supplementary Figure S7). We estimated the correlation of FGRS and the true genetic and environmental liability. For FGRS^21^ we included consider to versions: a c^2^-adjusted FGRS and an unadjusted FGRS. For PA-FGRS, we consider using all available relatives (PA-FGRS) or estimating liability *without parents, siblings, and children* (PA-FGRS_noFDR_) as a correction for shared environment. For each of the four (PA-)FGRS we computed the correlation between true and estimated liability in simulations.

### Psychiatric phenotypes

Our primary outcome, MDD, was defined as having a registration with a depressive episode (F32) or recurrent depression (F33) before Jan 1st 2017, according to the Danish Psychiatric Central Research Register (PCR)^36^. Diagnostic codes used for the construction of PA-FGRS scores are found in Supplementary Table S1. For relatives diagnosed between 1968 and 1994 records are limited to in-patient contacts and ICD-8 codes.

### Population parameters used for computing PA-FGRS in iPSYCH

The sex-specific lifetime prevalence of each disorder (Supplementary Table S1) was obtained from published estimates based on Danish registers^50^ . Narrow-sense heritability was set to 0.8 for ADHD, ASD, BPD and SCZ, and 0.4 for MDD (Supplementary Table S1). We chose to estimate K_i_ using sex and birth-year-specific cumulative incidence computed using all members of the iPSYCH2015 random sample genealogies (N=979,582; Supplementary Figure S14).

### Polygenic Scores

PGS for MDD, SCZ and BP were computed based on published, external summary statistics (Supplementary Table S2) that had no overlap with iPSYCH. PGS for ASD and ADHD were based on GWAS performed in the complementary half of the iPSYCH2015 case-cohort (i.e., iPSYCH2012 for iPSYCH2015i and vice versa, Supplementary Figure S9). We used SBayesR^51^ to estimate allelic effects for SNPs in the intersection of all GWAS, iPSYCH, and the reference LD panel. Palindromic SNPs (A/T, C/G), those not mapping uniquely to hg19 positions, and without a unique rsID in dbSNP v151 were excluded via our summary statistics QC pipeline (https://github.com/BioPsyk/cleansumstats).

### Classification analysis

In the European subset of the iPSYCH2015 MDD case-cohort (Supplementary Figure S9), we used logistic regression with MDD as an outcome and each or varying combinations of PA-FGRS and PGS as predictors (Supplementary Table S1-S2). For this analysis, PA-FGRS were computed excluding proband status. The classification accuracy was reported in an out-of-sample test. We trained the logistic classifier in iPSYCH2012 (or iPSYCH2015i) and reporting the area under the receiver operating characteristic curve (AUC) achieved in the independent, complementary iPSYCH2015i (or iPSYCH2012).

### Comparing Polygenic profiles

Putative subgroup-defining features were obtained from the PCR^36^ and the Danish Civil Registry^22^. We divided individuals diagnosed with MDD on the basis of a diagnosis of BPD (ICD10: F30-F31), comorbid anxiety (F40.0-40.2, F41.0-41.1, or F42), sex (as registered at birth), recurrence (ICD10: F32 or F33), severity (ICD10: F32/33.0, F32/33.1, F32/33.2, or F32/33.3), age at first recorded diagnosis, and mode of treatment (inpatient, casualty-ward or outpatient). We computed a composite estimate of genetic liability for each of the five mental disorders as a weighted sum of the PGS and PA-FGRS with weights corresponding to the betas from a logistic regression of their natural outcome in a calibration sample (Supplementary Figure S9). For each subgroup defining feature, multiple multinomial logistic regression was fitted to sequentially estimate the effects of each the composite genetic risk estimates with age, sex and 10 genetic PCs as covariates using the R package *nnet*^52^. We report a normalized partial effect size for each PGS and PA-FGRS, *β*_MLR_/*β*_LR_. The effect is the ratio of the effect of the PA-FGRS on MDD outcomes (*β*_MLR_) over its effect on the natural outcomes (*β*_LR_; e.g., ASD for FGRS for ASD).Each *β*_LR_ was estimated separately in outcome-specific case cohort samples (e.g. ASD case cohort, Supplementary Figure S9). This effect sizes can then be given context, for example, the effect of BPD genetic liability for being diagnosed BPD given a prior diagnosis of MDD is the same (*β*_MLR_/*β*_LR_ ∼ 1) as the effect of being diagnosed by BPD in the general population. These analyses were conducted separately for iPSYCH2012 and iPSYCH2015i samples and meta-analyzed. Subgroup-level effect estimates were meta-analyzed using inverse variance weighting, while heterogeneity test p–values were combined using Fisher’s method. In total we report 35 p-values declaring 0.05/35 = 0.0014 strictly significant.

### Genome-Wide Association Studies (GWAS)

GWAS were performed within two proband groups, the iPSYCH2012 MDD case-cohort and the iPSYCH2015i MDD case-cohort, on imputed allelic dosage data using plink2^53^. For binary MDD diagnosis, logistic regression was applied, for continuous valued PA-FGRS, we used linear regression, both including sex and age and 10 principal components of genetic ancestry as covariates. Inverse-variance weighted meta-analysis of the two constituent samples was performed using METAL^54^. SNPs with association p-values less than 5x10^-8^ were declared significant, while variants with a false discovery rate of 0.05 were considered suggestive. Independent loci were defined as >1Mb apart. Observed-scale SNP-heritability (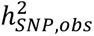) and genetic correlations to nine published GWAS (Supplementary Table S3) were estimated using LDscore regression^55, 56^. Difference in 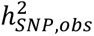 was computed as 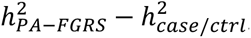, with std.err. 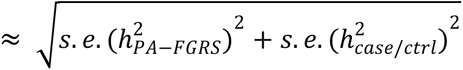. Genome-wide significant index SNPs were defined from a large external GWAS of MDD, modified to exclude 23andMe and iPSYCH, by clumping overlapping SNP lists. A paired t-test of the squared test statistic was used to assess significance of improvement. Polygenic scores for within iPSYCH classification were computed using SNPs with MAF>0.01 and INFO>0.8, clumped and thresholded with Plink 1.90b6.27^53^, using parameters --clump-kb 625 --clump-p1 0.1 --clump-p2 0.1 -- clump-r2 0.8. Improvements in predictions were assessed using the difference in AUC test in the pROC package.

## Data Availability

All data produced in the present study are available upon reasonable request to the authors and in accordance with Danish law.

