## Supplementary Figures for "PA-FGRS is a novel estimator of pedigree-based genetic liability that complements genotype-based inferences into the genetic architecture of major depressive disorder"

**Supplementary Figure S1 | Count of relative types in MDD case-cohort 2**
**Supplementary Figure S2 | Genotype-based estimates of kinship for reported full siblings, reported half siblings and siblings where information on at least one of the fathers was missing. 3**
**Supplementary Figure S3 | Genotype-based estimates of kinship for cousins whose parents are same sex twins, other cousins and half siblings. 4**
**Supplementary Figure S4 | Illustration of the PA-FGRS method 5**
**Supplementary Figure S5 | Calibration of FGRS in simulated pedigree data without censoring 6**
**Supplementary Figure S6 | Runtimes of the different methods for estimating genetic liabilities from phenotype data on genetic relatives 7**
**Supplementary Figure S7 | Effects of shared environment effects on the estimated genetic liabilities in simulated pedigree data. 8**
**Supplementary Figure S8 | Effect of mis-specified heritability or prevalence parameter in simulated pedigree data 9**
**Supplementary Figure S9 | Selection of the iPSYCH MDD case cohort. 10**
**Supplementary Figure 10 | Profiles of genetic liability to different mental disorders computed using PA-FGRS 11**
**Supplementary Figure 11 | Profiles of genetic liability to different mental disorders computed using PGS 12**
**Supplementary Figure 12 | Profiles of genetic liability to different mental disorders computed using PA-FGRS without first-degree relatives 13**
**Supplementary Figure 13 | Estimated power to detect an associated genetic variant using either logistic regression, or estimated genetic liability. 14**
**Supplementary Figure 14 | Cumulative incidence of five psychiatric disorders in the random population sample and their available relatives 15**

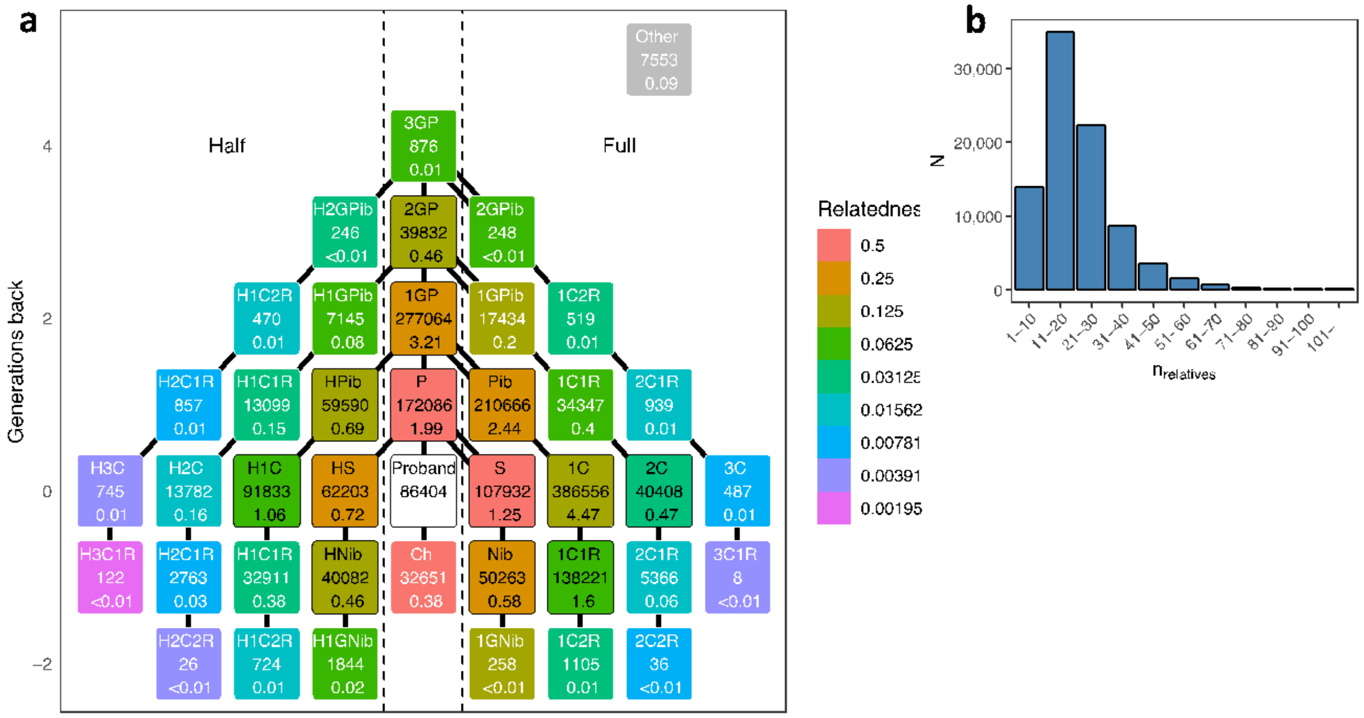


**Supplementary Figure S1 | Count of relative types in MDD case-cohort**

**a** Total and average per proband count of available relatives for the 86,404 individuals constituting the iPSYCH-2015 MDD case-cohort (**Proband**). **P** parents, **S** siblings, **Ch** children, **1GP** grandparents, **Pib**(lings) aunts and uncles, **Nib**(lings) nieces and nephews, **iCjR** $i^{th}$ cousin j removed, **H-** half, and **Other** relative types not in the figure. **b** Distribution of the number of relatives that could be linked to each of the 86,404 probands.

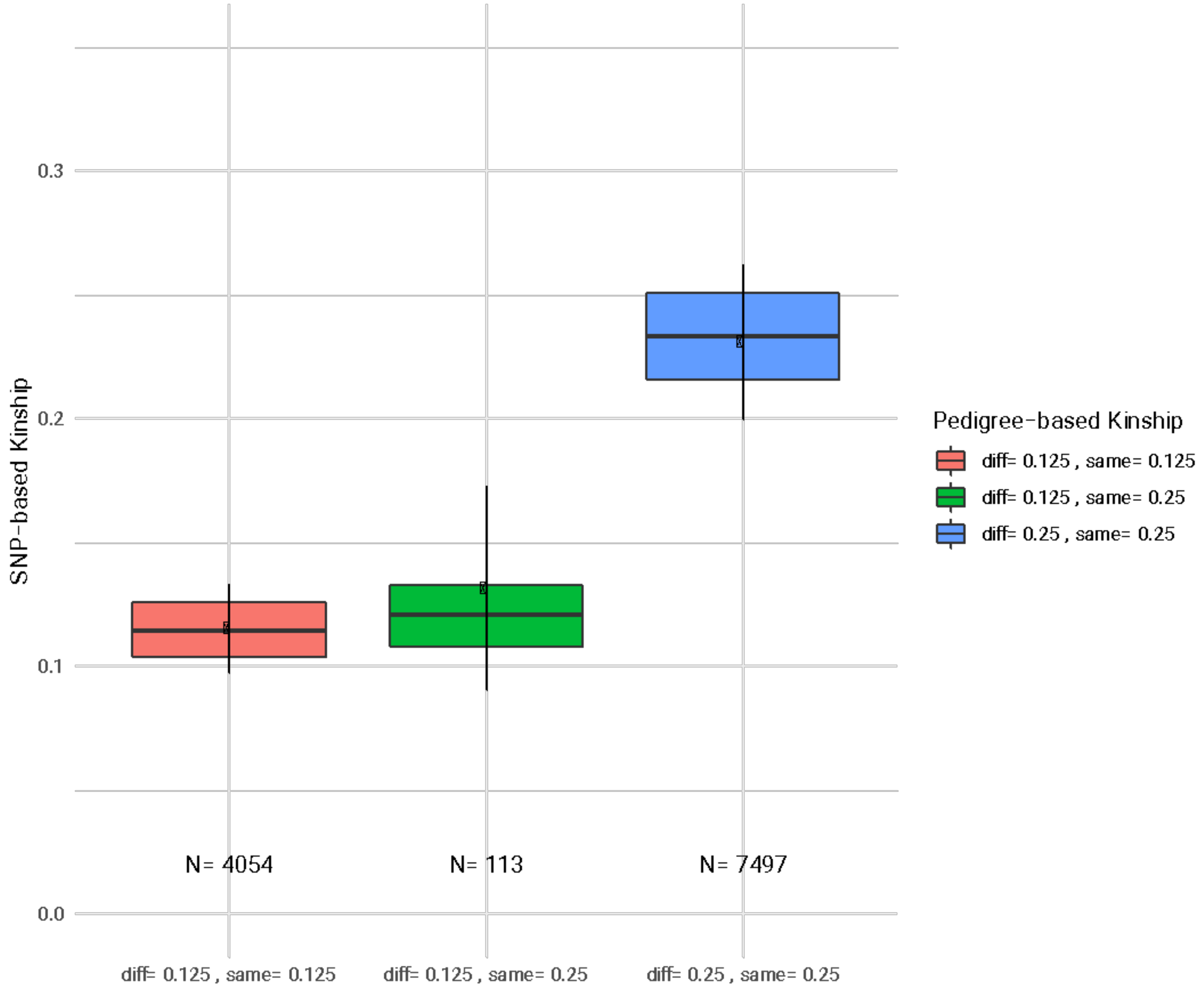


**Supplementary Figure S2 | Genotype-based estimates of kinship for reported full siblings, reported half siblings and siblings where information on at least one of the fathers was missing.**

We estimated the genotype-based relatedness of the 113 pairs of siblings where at least one father was missing. We found that the mean kinship was 0.14 and decided to treat all such cases as half siblings (relatedness = 0.25).

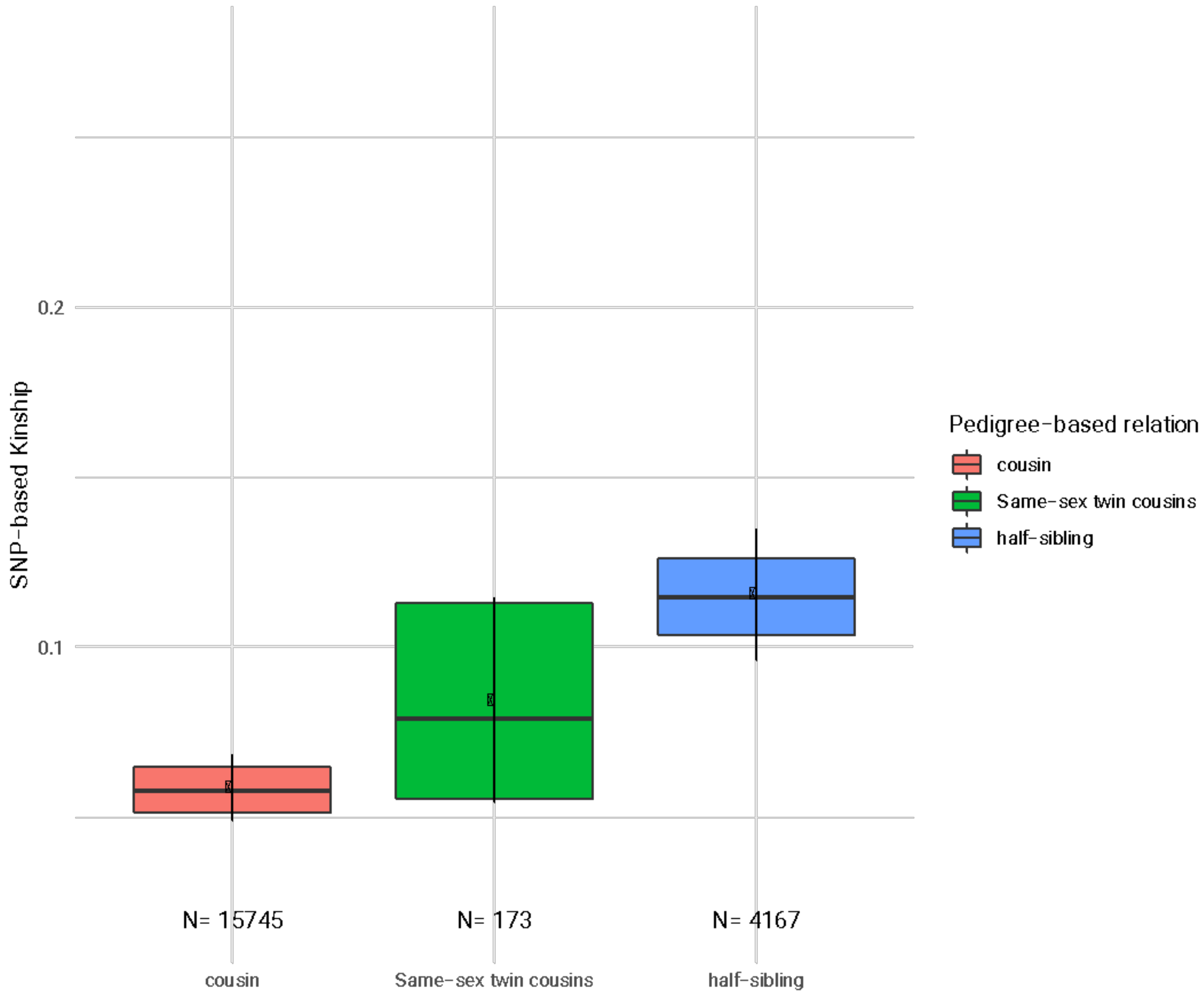


**Supplementary Figure S3 | Genotype-based estimates of kinship for cousins whose parents are same sex twins, other cousins and half siblings.**

We estimated genotype-based relatedness of the for 173 pairs of cousins for whom the parents were same-sex twins. We saw a mean kinship of 0.09 approximately half-way between other pairs of cousins and half-siblings, indicating that around half of the same-sex twins were monozygotic. We therefore decided to treat all same-sex twins as if they have relatedness of 0.75.

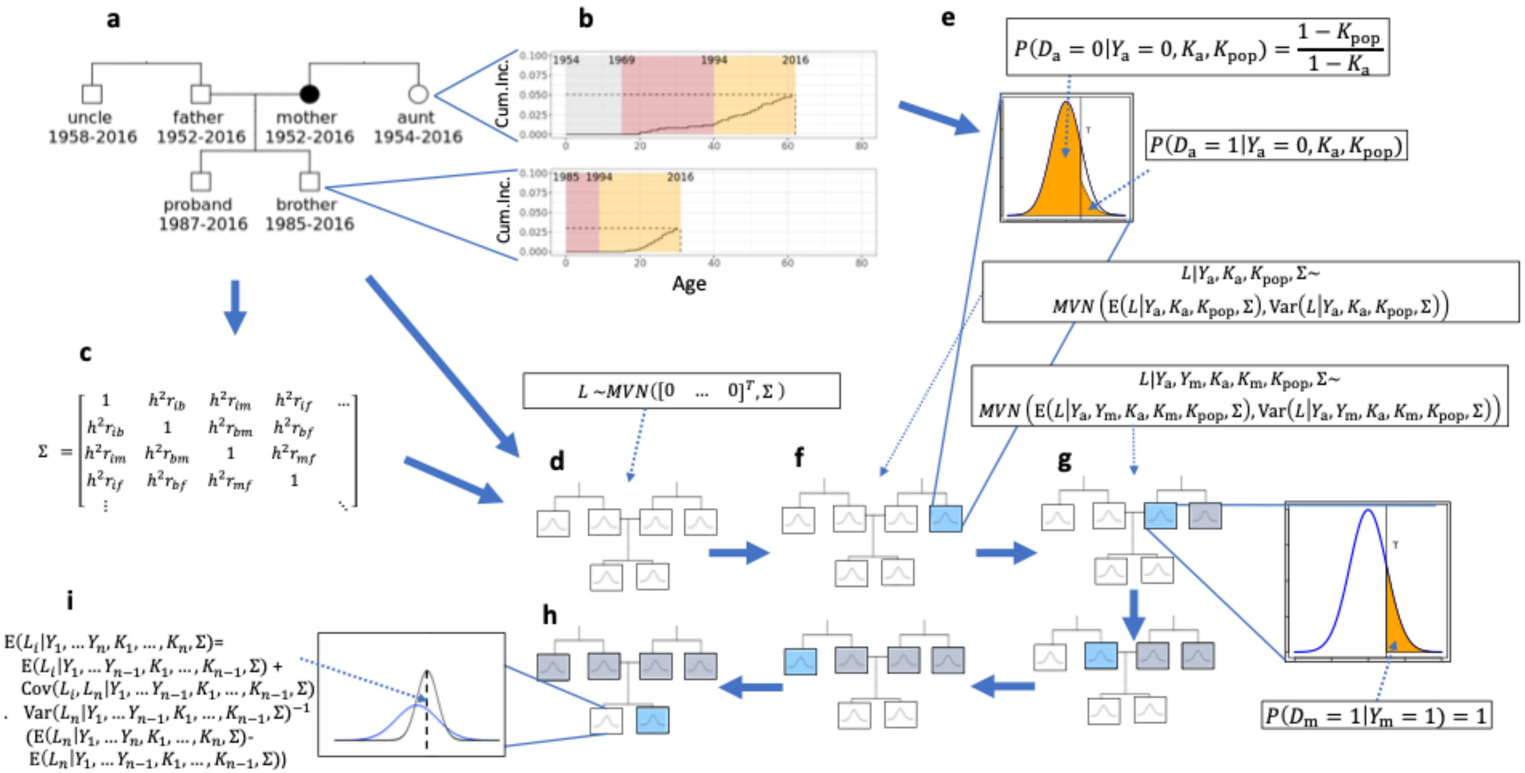


**Supplementary Figure S4 | Illustration of the PA-FGRS method**

**a**) Pedigree of a proband with one parent affected and four unaffected relatives. **b**) the cumulative incidence function of the brother and the aunt. **c**) Covariance of the liabilities given the kinship and an external estimate of the heritability. **d**) Marginal distributions of liabilities before conditioning on disease status. **e**) Distribution of liability of the aunt conditional on her disease status and the background population cumulative incidence at her age at the end of follow-up ($K_a$). **f**) Marginal distributions of liabilities after conditioning on disease status of the aunt. **g**) Marginal distributions of liabilities after conditioning on disease status of the aunt and the mother. **h**) Marginal distributions of liabilities after conditioning on disease status of all the relatives. **i)** Posterior mean liability of the proband conditioning on disease status of all the relatives.

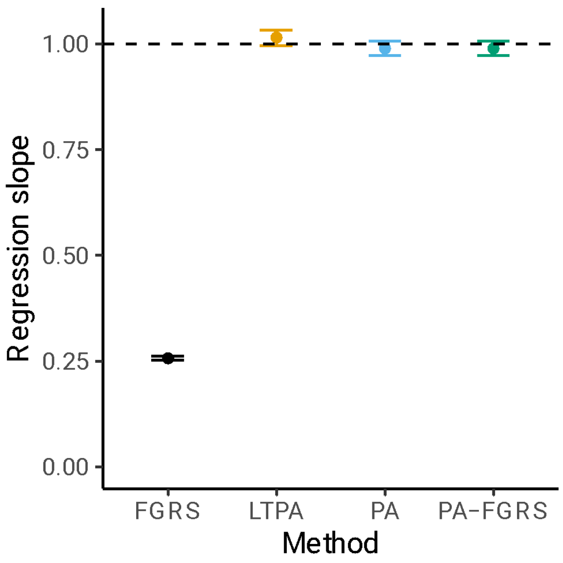


**Supplementary Figure S5 | Calibration of FGRS in simulated pedigree data without censoring**
Calibration of the FGRS estimated by four different methods in simulations without censoring, assessed by estimating the change in the true liability per unit change in the estimated liability by a linear regression with true liability as dependent variable. **FGRS** indicates the method proposed by Kendler et al., **LTPA** indicates the method by Hujoel et al., **PA** indicates the method by So et al., and **PA-FGRS** indicates the method proposed in this paper. Reported distributions are based on 100 simulations of 1000 pedigrees with $h^2$=0.5 and prevalence=0.40

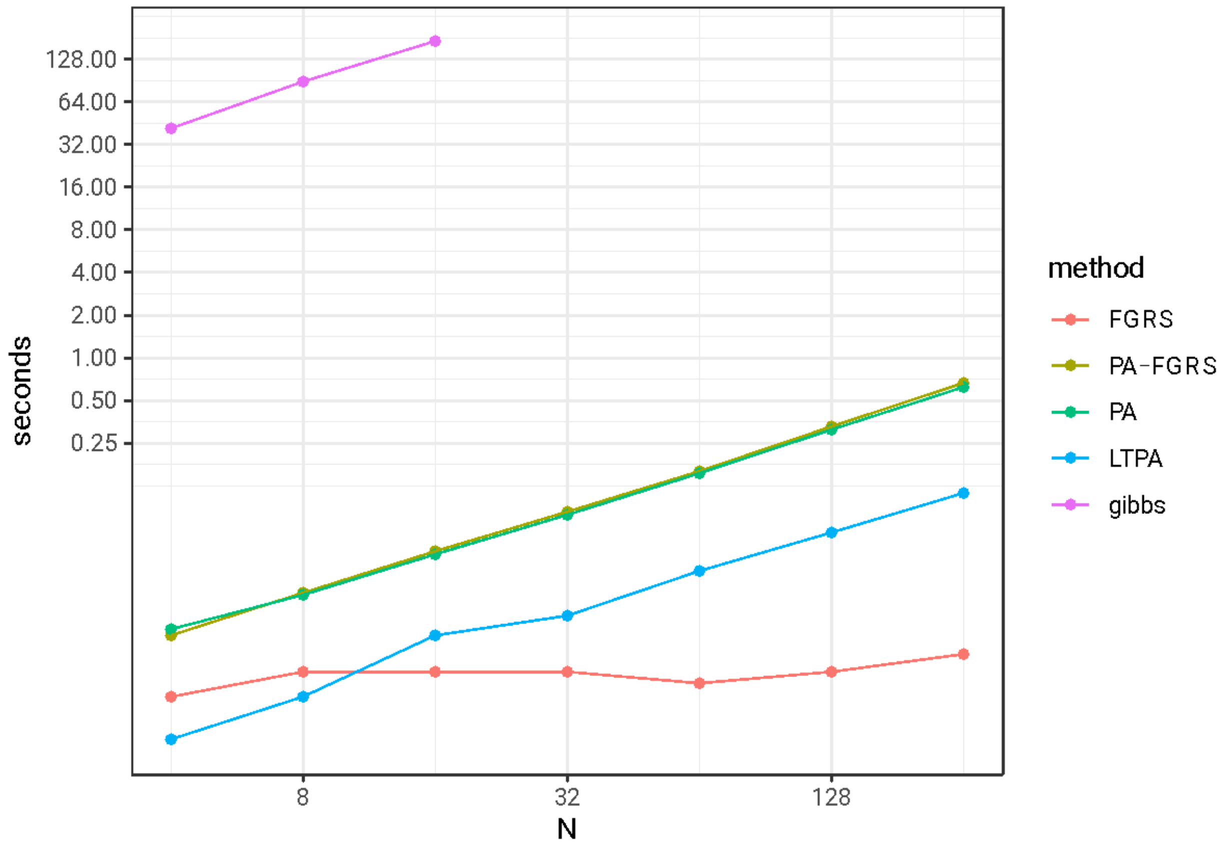


**Supplementary Figure S6 | Runtimes of the different methods for estimating genetic liabilities from phenotype data on genetic relatives**

Computation time for the five different estimators of genetic liability. **FGRS** indicates the method proposed by Kendler et al., **LTPA** indicates the method by Hujoel et al., **PA** indicates the method by So et al., and **PA-FGRS** indicates the method proposed in this paper.

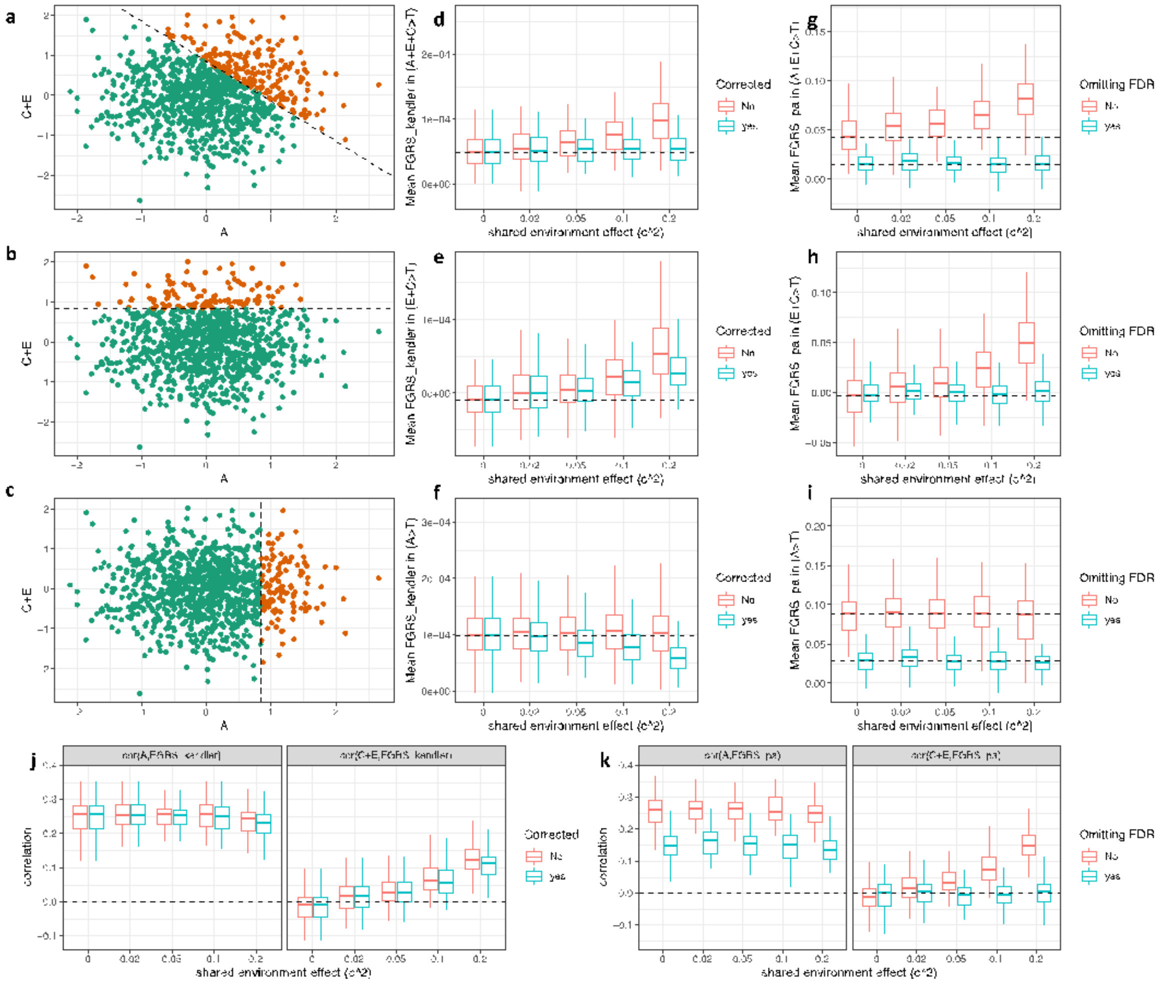


**Supplementary Figure S7 | Effects of shared environment effects on the estimated genetic liabilities in simulated pedigree data.**

Here we display the effect of shared environment effects, which we model by letting the liability of each individual be the sum of an additive genetic effect (**A**) and an environmental effect (**C+E**). **A** has a covariance matrix defined by the heritability and the relatedness, and **C+E** has a covariance matrix where the diagonal elements are (1-$h^2$) and the off-diagonal elements of first-degree relatives are ($c^2$) while the other off-diagonal elements are set to 0.

Panels **a-c** illustrates three different subset of the population, where **a** is the case-control definition under a liability-threshold ACE model, while **b** and **c**, illustrates subsets bases on either A only or C+E only being above a threshold, T. Panels **d-f** shows how the mean estimate of liability in cases by $FGRS_{Kendler}$, with and without correction for shared environment, under these three types subpopulation definitions, and under increasing levels of $c^2$. While the $c^2$-correction used by $FGRS_{Kendler}$ has the effect of making the mean score of cases independent of the level of $c^2$ (**d**), but does not prevent that the mean score in the subpopulation with C+E>T (**b**) increases with increasing levels of $c^2$. Our proposed correction to PA-FGRS (**g-i**) makes estimated scores independent of the level of $c^2$. Panels **j-k** shows the estimated correlations between the estimated liability and true value of **A** and **C+E** for $FGRS_{Kendler}$ with and without correction (**j**) for and PA-FGRS including or omitting first degree relatives (**k**)**.** In summary our simulations show that while the correction proposed in $FGRS_{Kendler}$ gives unbiased estimates of liability in cases, it is not uncorrelated with C, this can be obtained by estimating lability only from non-first degree relatives, but this lowers the correlation with A. Reported distributions in panels **d-k** are based on 100 simulations of 500 pedigrees with $h^2$=0.5 and prevalence=0.2.

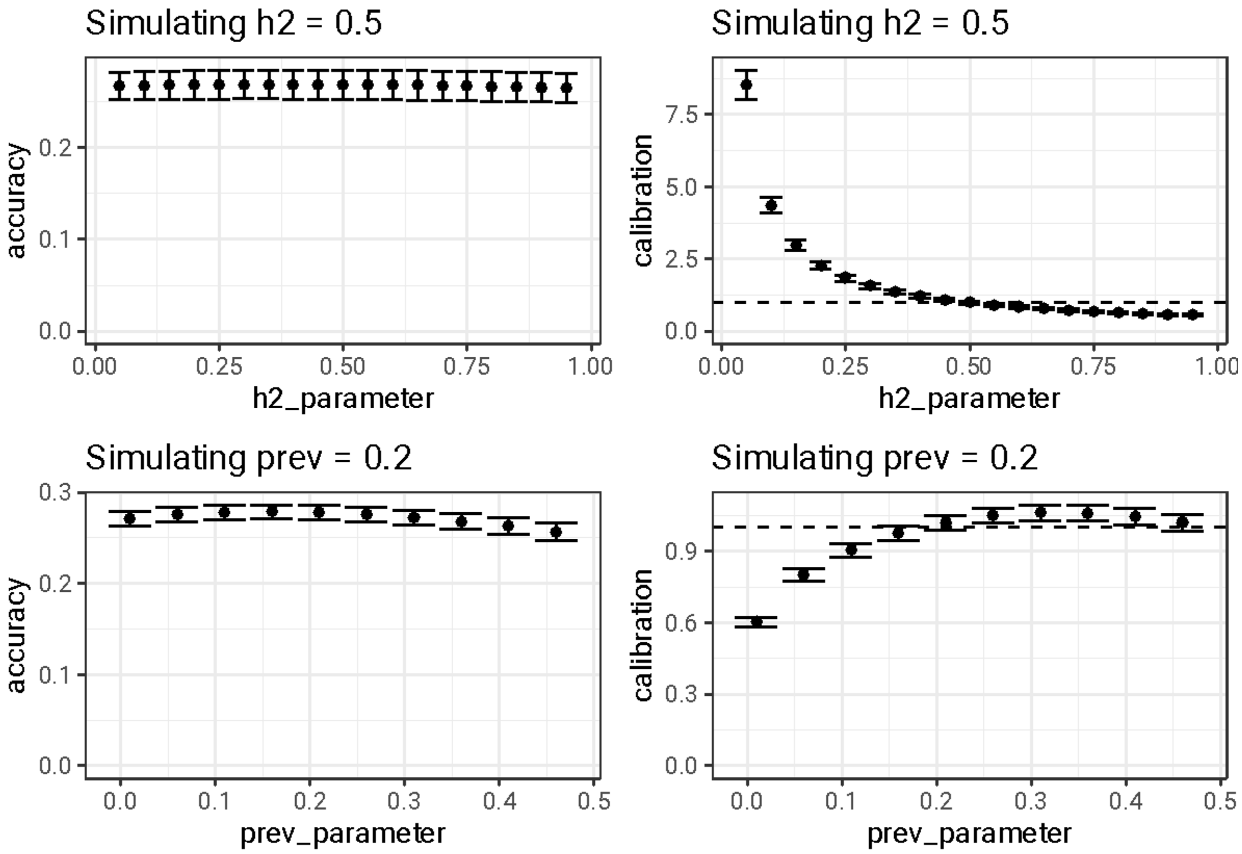


**Supplementary Figure S8 | Effect of mis-specified heritability or prevalence parameter in simulated pedigree data**

The figure displays the effect of simulating pedigree data with a true heritability of 0.5 and prevalence of 0.2 and then running the PA-FGRS with a range of different heritability or parameters. In the left panels, we display the distribution estimated correlation with the true genetic liability obtained across simulations. In the right panels, we display the distribution of the estimated increase in true liability per unit increase in the estimated liability (a measure of calibration) across simulations. The reported distributions are based on 10 simulations of 500 pedigrees.

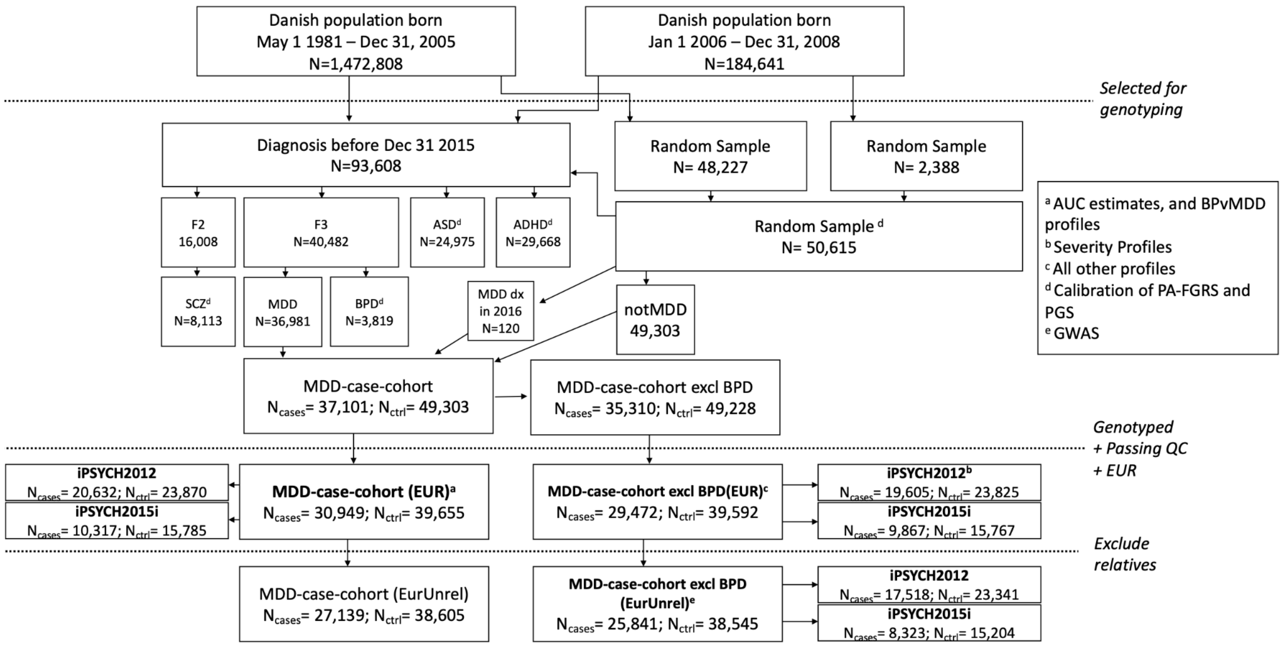


**Supplementary Figure S9 | Selection of the iPSYCH MDD case cohort.**
Diagram showing the selection of the iPSYCH-2015 case cohort, and the different subcohort used in this study from the Danish birth cohort.

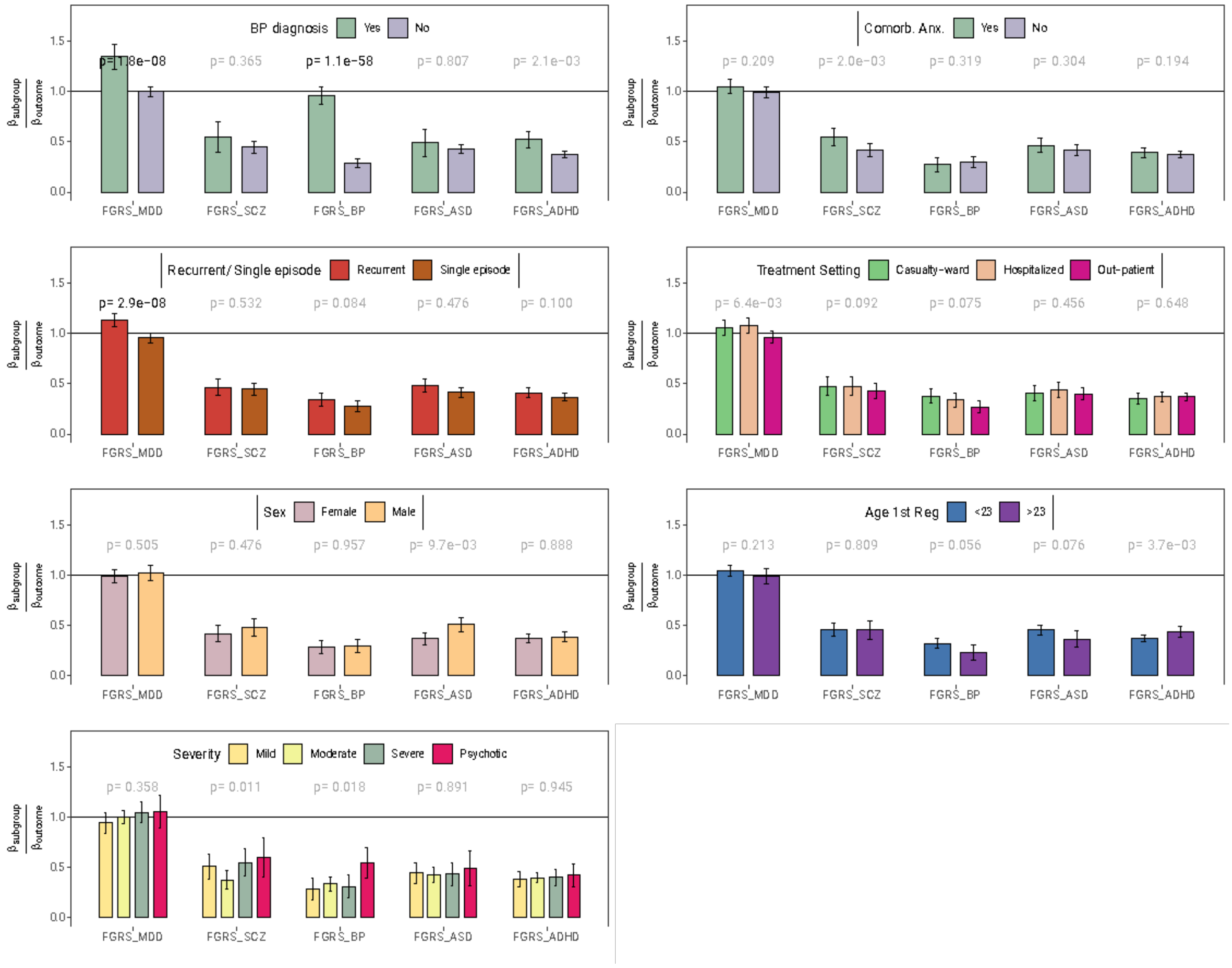


**Supplementary Figure 10 | Profiles of genetic liability to different mental disorders computed using PA-FGRS**

The figure shows the results of a multinomial logistic regression where the dependent variable is the categories of no-diagnosis and (**a**) MDD with/without bipolar disorder diagnosis, (**b**) MDD with/without comorbid anxiety, (**c**) single depressive episode and recurrent MDD (**d**) out-patient, casualty-ward and inpatient diagnoses of MDD (**e**) female MDD and male MDD, (**f**) first MDD diagnosis before/after age 23, (**g**), mild, moderate, severe, or psychotic depression. The independent variable is a composite estimate of genetic risk of one of five different mental disorders, constructed as a weighted sum of PA-FGRS and PGS. The y-axis indicates the estimated coefficient divided by the coefficient for the target diagnosis in a binomial logistic regression. **MDD** major depressive disorder, **SCZ** schizophrenia, **BPD** bipolar disorder, **ASD** autism spectrum disorders, **ADHD** attention-deficit/ hyperactivity disorder**, p** the probability of observing this data under the null-hypothesis (that all outcomes have the same coefficient). P-value in black indicates $p<0.05/35$. Beta-estimates and p-values are meta-analyzed across iPSYCH-2012 ( $N_{cases} \leq 20{,}632$, $N_{ctrl} \leq 23{,}870$) and iPSYCH2015i ($N_{cases} \leq 10{,}317$, $N_{ctrl} \leq 15{,}785$) with the exception of panel **g** (Severity) which are from iPSYCH2012 only. Sample sizes for the individual analyses are provided in Table S3. Error-bars indicate 95% confidence intervals.

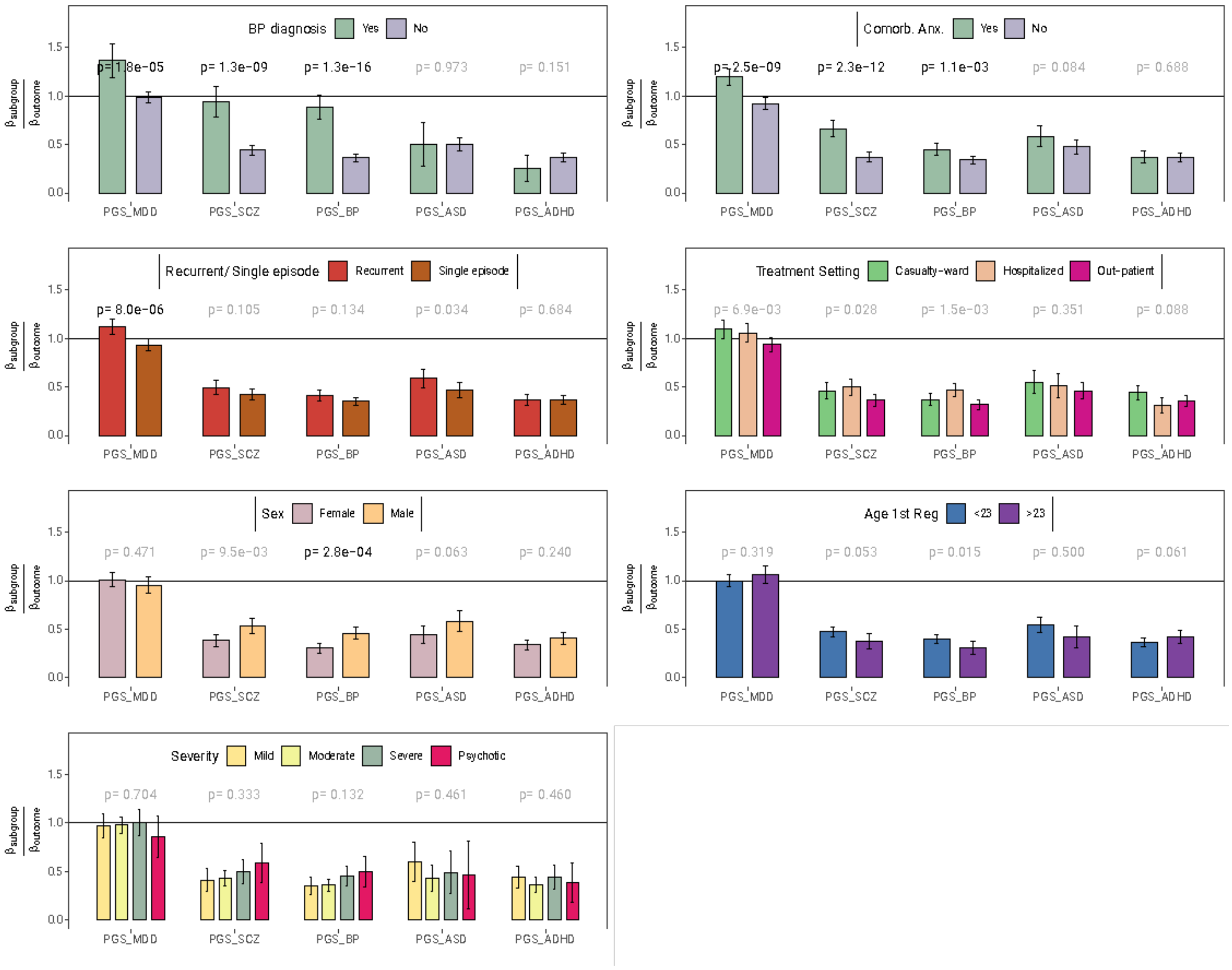


**Supplementary Figure 11 | Profiles of genetic liability to different mental disorders computed using PGS**
The figure shows the results of a multinomial logistic regression where the dependent variable is the categories of no-diagnosis and (a) MDD with/without bipolar disorder diagnosis, (b) MDD with/without comorbid anxiety, (c) single depressive episode and recurrent MDD (d) out-patient, casualty-ward and inpatient diagnoses of MDD (e) female MDD and male MDD, (f) first MDD diagnosis before/after age 23, (g), mild, moderate, severe, or psychotic depression. The independent variable is a composite estimate of genetic risk of one of five different mental disorders, constructed as a weighted sum of PA-FGRS and PGS. The y-axis indicates the estimated coefficient divided by the coefficient for the target diagnosis in a binomial logistic regression. MDD major depressive disorder, SCZ schizophrenia, BPD bipolar disorder, ASD autism spectrum disorders, ADHD attention-deficit/ hyperactivity disorder, p the probability of observing this data under the null-hypothesis (that all outcomes have the same coefficient). P-value in black indicates $p<0.05/35$. Beta-estimates and p-values are meta-analyzed across iPSYCH-2012 ( $N_{cases}\leq 20{,}632$, $N_{ctrl}\leq 23{,}870$) and iPSYCH2015i ($N_{cases}\leq 10{,}317$, $N_{ctrl}\leq 15{,}785$) with the exception of panel g (Severity) which are from iPSYCH2012 only. Sample sizes for the individual analyses are provided in Table S3. Error-bars indicate 95% confidence intervals.

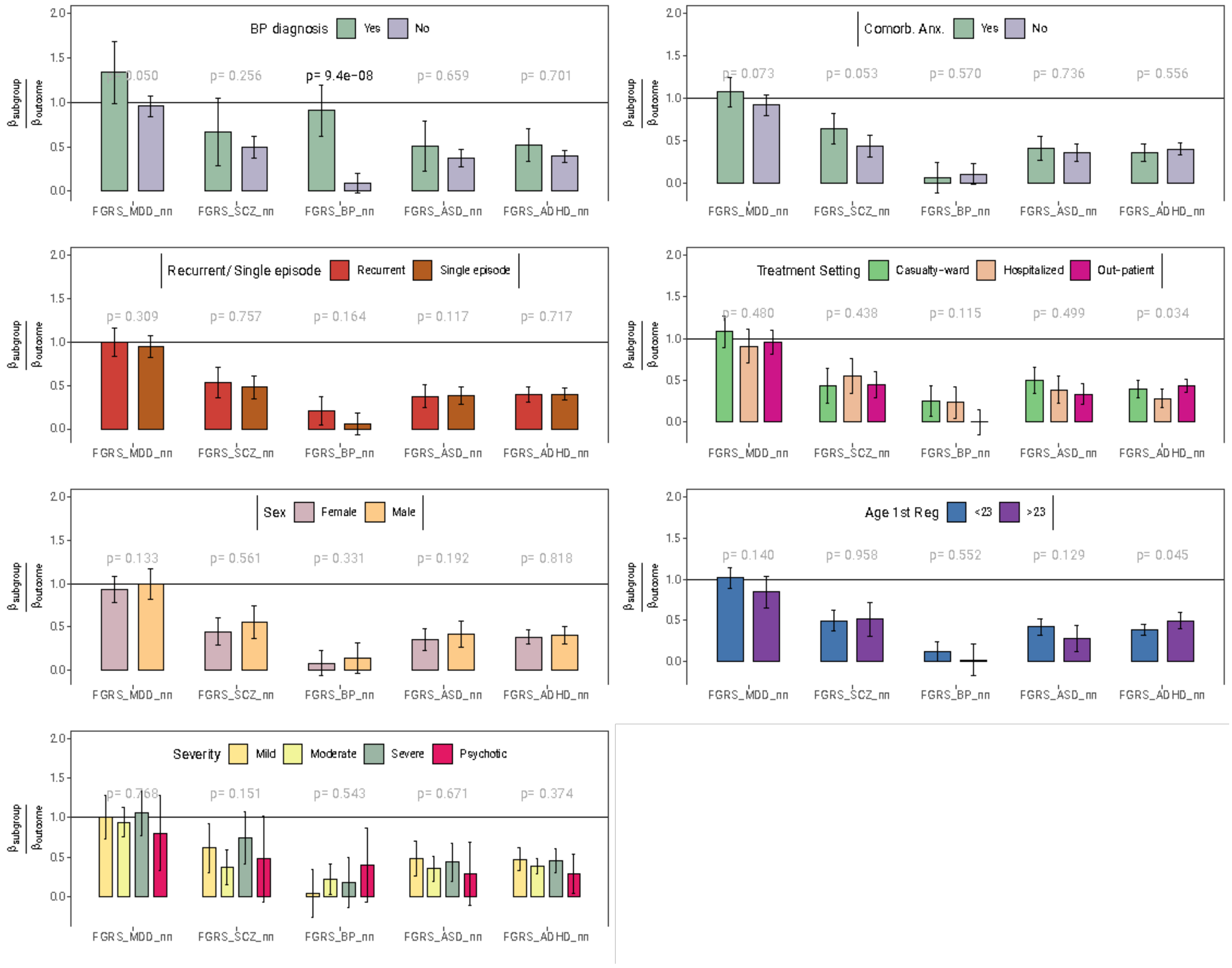


**Supplementary Figure 12 | Profiles of genetic liability to different mental disorders computed using PA-FGRS without first-degree relatives**

The figure shows the results of a multinomial logistic regression where the dependent variable is the categories of no-diagnosis and (**a**) MDD with/without bipolar disorder diagnosis, (**b**) MDD with/without comorbid anxiety, (**c**) single depressive episode and recurrent MDD (**d**) out-patient, casualty-ward and inpatient diagnoses of MDD (**e**) female MDD and male MDD, (**f**) first MDD diagnosis before/after age 23, (**g**), mild, moderate, severe, or psychotic depression. The independent variable is a composite estimate of genetic risk of one of five different mental disorders, constructed as a weighted sum of PA-FGRS and PGS. The y-axis indicates the estimated coefficient divided by the coefficient for the target diagnosis in a binomial logistic regression. **MDD** major depressive disorder, **SCZ** schizophrenia, **BPD** bipolar disorder, **ASD** autism spectrum disorders, **ADHD** attention-deficit/ hyperactivity disorder**, p** the probability of observing this data under the null-hypothesis (that all outcomes have the same coefficient). P-value in black indicates $p<0.05/35$. Beta-estimates and p-values are meta-analyzed across iPSYCH-2012 ( $N_{cases}$≤20,632, $N_{ctrl}$≤23,870) and iPSYCH2015i ($N_{cases}$≤10,317, $N_{ctrl}$≤15,785) with the exception of panel **g** (Severity) which are from iPSYCH2012 only. Sample sizes for the individual analyses are provided in Table S3. Error-bars indicate 95% confidence intervals.

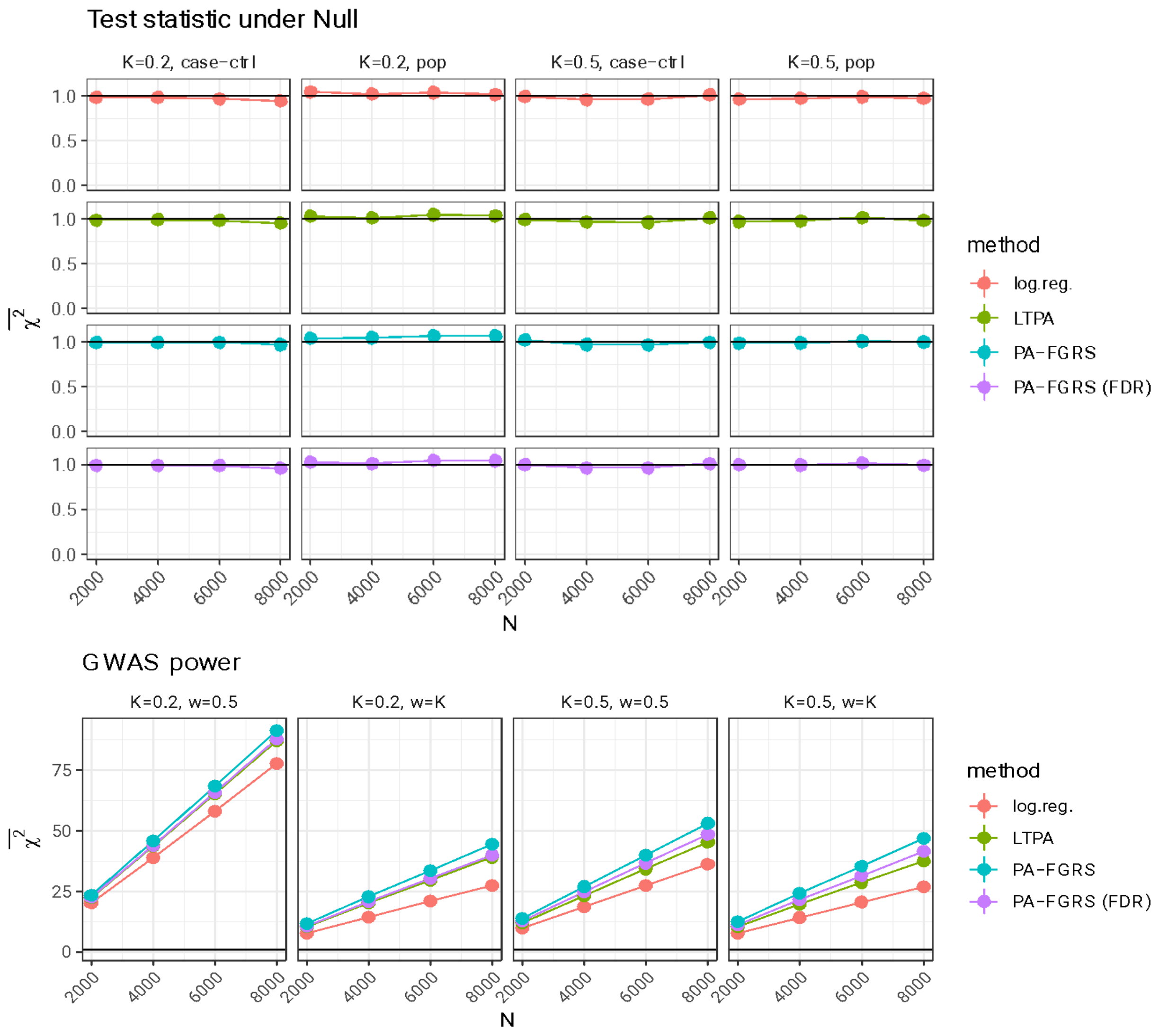


**Supplementary Figure 13 | Estimated power to detect an associated genetic variant using either logistic regression, or estimated genetic liability.**

To assess the power to detect a genetic variant associated with the liability to disease, we simulate 50,000 pedigrees with (0-18 relatives) where the 20 independent normally distributed variables each explain 5% of the variance in the genetic liability and 20 random gaussian variables (null). We generate phenotypes according to a liability threshold model with prevalence (K) of either 0.2 or 0.5, $h^2$=0.5, and cumulative incidence curve linearly increasing from age 10 to age 65. From these, we sample a number individuals (N) either as population sampling (w=K) or half cases, half controls (w=0.5). Based on the observed disease status and family history ( We then run association tests by four different methods: (1) as logistic regression on proband status (LR); (2) as a linear regression on estimated liabilities based on disease status and parents and sibling history using the method by Hujoel et al. (LTPA); (3) as a linear regression on estimated liabilities based on disease status and parents and sibling history using the method proposed in this paper (PA-FGRS (FDR)); and (4) as a linear regression on estimated liabilities based on disease status and all family history using the method proposed in this paper (PA-FGRS). This is repeated 100 times and and the mean $\chi^2$for reported for null variants (above) and causal variants (below).

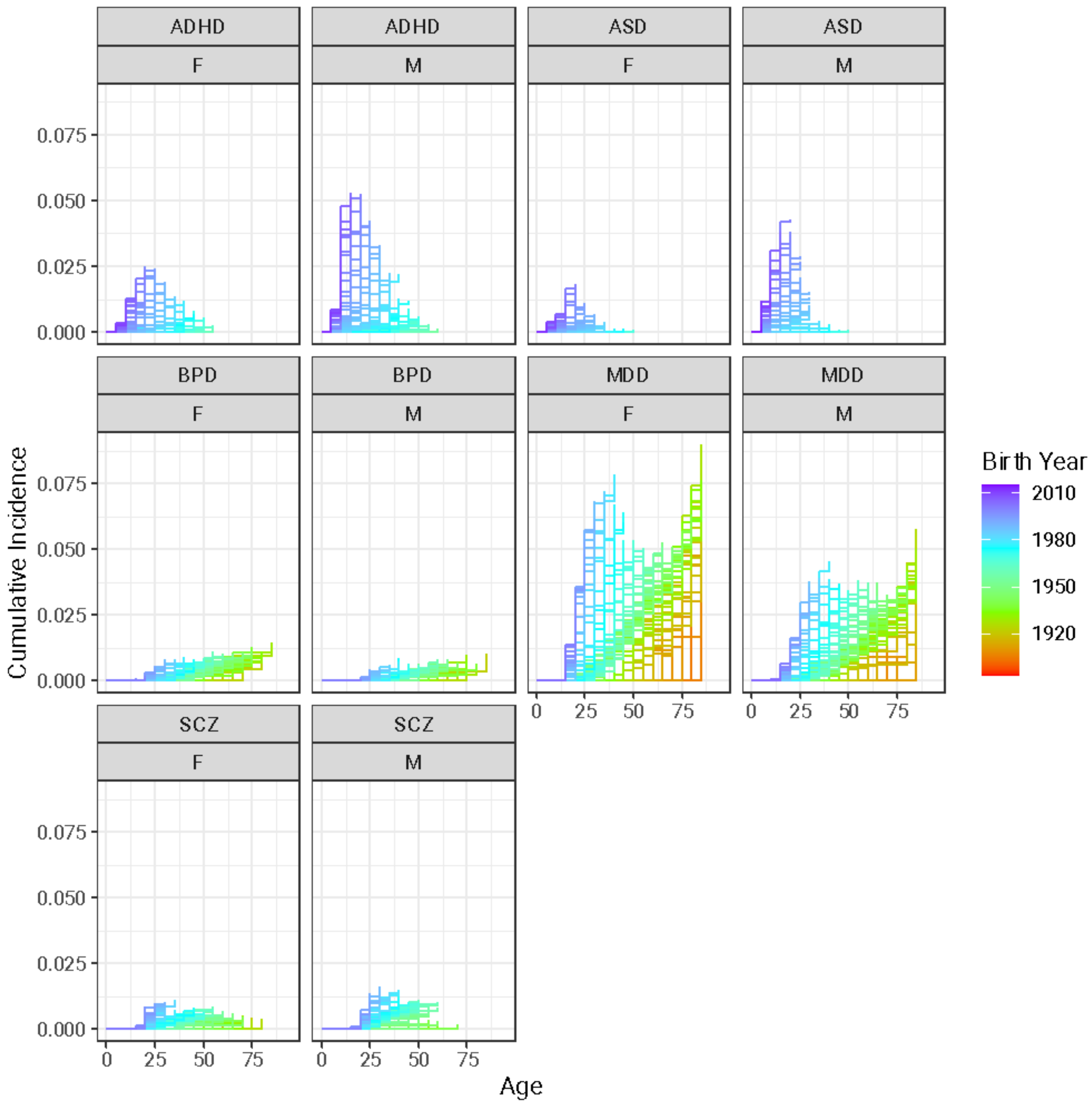


**Supplementary Figure 14 | Cumulative incidence of five psychiatric disorders in the random population sample and their available relatives**

Estimated cumulative incidence of registered diagnoses of ADHD, ASD, BPD, MDD and schizophrenia estimated separately for females (F) and males (M), a stratified by year of birth (color). For visualization, the cumulative incidence curves simplified such that a step, corresponds to minimum five new diagnosed cases and time is is measured in five-year increments.
