## Supplementary Information for "PA-FGRS is a novel estimator of pedigree-based genetic liability that complements genotype-based inferences into the genetic architecture of major depressive disorder"

**Appendix**

The Pearson-Aitken Family Genetic Risk Score (PA-FGRS) is a novel estimator for the expected genetic liability to disease carried by a proband given the pattern of diseases in an arbitrarily structured pedigree of relatives that may be only partially observed. This estimator is derived under a modified version of the liability threshold model for disease [1]. The method first estimates an initial liability for each relative and then uses the Pearson-Aitken selection formula to sequentially update the expected liability in the proband conditional on each relative.[1,2]

First, as outlined in the main text, we assume that a disease $D_i$ is defined as:

$$D_i = \begin{cases} 1 & \text{if } L_i > t \\ 0 & \text{if } L_i < t \end{cases} \qquad L_i = \sum_j \beta_j X_{ij} + e_i$$

Where $L \sim N(0, 1)$ such that the prevalence in the population $K_{pop}$ is given by the cumulative distribution function of the standard normal distribution $K_{pop} = 1 - \Phi(T)$.
However, if the disease has age of onset later than at birth, the observed disease status $Y_i$ can be different from $D_i$ (i.e. $Y \leq D$), when an individual $i$ has only been observed for a fraction of the risk window. We can express this as:

$$Y_i = \begin{cases} \text{Bernoulli}(\frac{K_i}{K_{\text{pop}}}) & \text{if } D_i = 1 \\ 0 & \text{if } D_i = 0 \end{cases} \qquad \text{(Eq. S1)}$$

Where $K_i$ is the prevalence of the disease at age of individual $i$ and $K_{pop}$ is the population lifetime prevalence of the disease. Our aim is to estimate the expected liability of an individual given the observed disease status of a number of relatives and their age at the end of follow-up.

**Covariance matrix of liabilities**

For an individual $p$ with $n$ relatives, we assume that the covariance matrix ($\Sigma$) of the random vector of liabilities, $L = \left[L_1, \ldots, L_n, L_p, G_p\right]^T$, is given by the heritability of the phenotype ($h^2$) and the ($n$+2 by $n$+2) genetic relatedness matrix, $r$, such that:

$$\Sigma = \begin{bmatrix} 1 & \cdots & r_{1,n}h^2 & r_{1,p}h^2 & r_{1,p}h^2 \\ \vdots & \ddots & \vdots & \vdots & \vdots \\ r_{n,1}h^2 & \cdots & 1 & r_{n,p}h^2 & r_{n,p}h^2 \\ r_{p,1}h^2 & \cdots & r_{p,n}h^2 & 1 & h^2 \\ r_{p,1}h^2 & \cdots & r_{p,n}h^2 & h^2 & h^2 \end{bmatrix}$$

**Conditional liability distribution:**

Let $\mu^{i*}$ and $\Omega^{i*}$ denote the expected mean vector and variance covariance matrix of $L$ conditional on disease status of relatives $1, \ldots, i$. Such that:

$$\mu_j^{i*} = E(L_j | Y_1, \ldots, Y_i, K_1, \ldots, K_i, K_{pop}, \Sigma)$$
$$\Omega_{j,k}^{i*} = Cov(L_j, L_k | Y_1, \ldots, Y_i, K_1, \ldots, K_i, K_{pop}, \Sigma)$$

The algorithm we use to estimate genetic liability works by conditioning on each of the relatives in turn. We start by setting the $\mu^{0*} = [0, \ldots, 0]^T$ and $\Omega^{0*} = \Sigma$. Below we derive the necessary steps for estimating the conditional liabilities.

We assume that the posterior liability of the $i^{th}$ individual is a mixture of two truncated normal distributions:

$$L_i | Y_1, \ldots, Y_i, K_1, \ldots K_i, K_{pop}, \Sigma \sim (1 - \pi_i)\psi(\mu_i^{(i-1)*}, \Omega_{i,i}^{(i-1)*}, a = -\infty, b = T) + \pi\ \psi(\mu_i^{(i-1)*}, \Omega_{i,i}^{(i-1)*}, a = T, b = \infty)$$

(Eq. S2)

where $\psi(\mu\ , \sigma\ , a, b)$ denotes a truncated normal normal distribution with left-truncation at $a$ and right-truncation at $b$.
Note that while this (Eq. S2) is strictly true for $i = 1$, it is only an approximation for $i > 1$, since the conditional distributions are no longer exactly Gaussian. [3] However, unless the disease is rare and the off-diagonal elements of $\Sigma$ are high (which will only occur for high $h^2$ in MZ-twins[3]) the deviation from normality is minor. We refer to this assumption as *conditional normality.*

**Deriving the mixture parameter**

The mixture parameter $\pi_i$ is given by the probability of being a case conditional on observed disease status of relatives 1 to i: $\pi_i = P(D_i = 0 | Y_1, \ldots, Y_{i-1}, Y_i = 0, K_1, \ldots, K_i, K_{pop}, \Sigma)$ . In the case of $Y_i = 1$ we have $\pi_i = 1$, whereas in the case $Y_i = 0$, $\pi_i$ is given by:

$$1 - \pi_i = P(D_i = 0 | Y_1, \ldots, Y_{i-1}, Y_i = 0, K_1, \ldots, K_i, K_{pop}, \Sigma)$$

by Bayes' theorem we have:

$$1 - \pi_i = \frac{P(D_i = 0 | Y_1, \ldots, Y_{i-1}, K_1, \ldots, K_i, K_{pop}, \Sigma) P(Y_i = 0 | D_i = 0, Y_1, \ldots, Y_{i-1}, K_1, \ldots, K_i, K_{pop}, \Sigma)}{P(Y_i = 0 | Y_1, \ldots, Y_{i-1}, K_1, \ldots, K_i, K_{pop}, \Sigma)}$$

and since $P(Y_i = 0 | D_i = 0, Y_1, \ldots, Y_{i-1}, Y_i = 0, K_1, \ldots, K_i, K_{pop}, \Sigma) = 1$

$$1 - \pi_i = \frac{P(D_i=0|Y_1,...,Y_{i-1},K_1,...,K_i,K_{pop},\Sigma)}{P(Y_i=0|Y_1,...,Y_{i-1},K_1,...,K_i,K_{pop},\Sigma)}$$

From our assumption that $P(Y_i = 1|D_i = 1)$ depends only on $K_i$ and $K_{pop}$ (Eq.S1), it follows that $P(Y_i = 1|D_i = 1, Y_1,..., Y_{i-1}, K_1,..., K_i, K_{pop}, \Sigma) = P(Y_i = 1|D_i = 1, K_i, K_{pop}) = \frac{K_i}{K_{pop}}$.

We can use this and the law of total probability to split up the denominator:

$$1 - \pi_i = \frac{P(D_i=0|Y_1,...,Y_{i-1},K_1,...,K_i,K_{pop},\Sigma)}{P(D_i=0|Y_1,...,Y_{i-1},K_1,...,K_i,K_{pop},\Sigma)+\left(1-\frac{K_i}{K_{pop}}\right)P(D_i=1|Y_1,...,Y_{i-1},K_1,...,K_i,K_{pop},\Sigma)}$$

$$= \frac{P(L_i<T|\mu_i^{(i-1)*},\Omega_{i,i}^{(i-1)*},T)}{P(L_i<T|\mu_i^{(i-1)*},\Omega_{i,i}^{(i-1)*},T)+\frac{K_{pop}-K_i}{K_{pop}}P(L_i>T|\mu_i^{(i-1)*},\Omega_{i,i}^{(i-1)*},T)}$$

assuming *conditional normality* we can replace the probabilities with the cumulative distribution function of the normal distribution:

$$1 - \pi_i \approx \frac{\Phi(\frac{T-\mu_i^{(i-1)*}}{\sqrt{\Omega_{i,i}^{(i-1)*}}})}{\Phi(\frac{T-\mu_i^{(i-1)*}}{\sqrt{\Omega_{i,i}^{(i-1)*}}})+\frac{K_{pop}-K_n}{K_{pop}}(1-\Phi(\frac{T-\mu_i^{(i-1)*}}{\sqrt{\Omega_{i,i}^{(i-1)*}}}))} \quad \text{(Eq.S3)}$$

**Expected liability conditional on disease status**

Having derived the mixture parameter ($\pi_i$), we can obtain the expected value $L_i$ conditional on $Y_1,..., Y_i$ and $K_1, \ldots K_i$ as the expected value of the mixture distribution:

$$\mu_i^{i*} = E(L_i|Y_1,..., Y_i, K_1, \ldots K_i, K_{pop}, \Sigma) = \pi_i E(L_i|L_i > T, \mu_i, \Omega_{i,i}, T) + (1 - \pi_i)E(L_i|L_i < T, \mu_i, \Omega_{i,i}, T)$$

assuming *conditional normality*, and inserting the expected value of a truncated normal, this can be approximated by:

$$\mu_i^{i*} \approx \pi_i(\mu_i^{(i-1)*} + \sqrt{\Omega_{i,i}^{(i-1)*}}\frac{\phi(\lambda)}{1-\Phi(\lambda)}) + (1 - \pi_i)(\mu_i^{(i-1)*} - \sqrt{\Omega_{i,i}^{(i-1)*}}\frac{\phi(\lambda)}{\Phi(\lambda)}) \quad \text{(Eq.S4)}$$

where $\lambda = \frac{T-\mu_i^{(i-1)*}}{\sqrt{\Omega_{i,i}^{(i-1)*}}}$ and with $\phi$ and $\Phi$ denoting the probability density function and the cumulative distribution function of the standard normal distribution.

**Expected variance conditional on disease status**

The expected variance of $L_i$ conditional on $Y_1,..., Y_i$ and $K_1, \dots K_i$ is obtained as the expected variance of the mixture distribution which is given by:

$$\Omega^{i*}_{i,i} = \mathrm{Var}(L_i | Y_1,..., Y_i, K_1, \dots K_i, K_{pop}, \Sigma) = \\ \pi_i (E(L_i | L_i > T, \mu_i^{(i-1)*}, \Omega_{i,i}^{(i-1)*}, T)^2 + \mathrm{Var}(L_i | L_i > T, \mu_i^{(i-1)*}, \Omega_{i,i}^{(i-1)*}, T) ) + \\ (1 - \pi_i)(E(L_i | L_i < T, \mu_i^{(i-1)*}, \Omega_{i,i}^{(i-1)*}, T)^2 + \mathrm{Var}(L_i | L_i < T, \mu_i^{(i-1)*}, \Omega_{i,i}^{(i-1)*}, T) ) - \\ \left(\pi_i E(L_i | L_i > T, \mu_i^{(i-1)*}, \Omega_{i,i}^{(i-1)*}, T) + (1 - \pi_i) E(L_i | L_i < T, \mu_i^{(i-1)*}, \Omega_{i,i}^{(i-1)*}, T)\right)^2$$

assuming *conditional normality*, and inserting the expected variance of a the right and left truncated normal distribution, this can be approximated by:

$$\Omega^{i*}_{i,i} \approx \pi_i \left( \left( \mu_i^{(i-1)*} + \sqrt{\Omega_{i,i}^{(i-1)*}} \frac{\phi(\lambda)}{1-\Phi(\lambda)} \right)^2 + \Omega_{i,i}^{(i-1)*} \left( 1 + \lambda \frac{\phi(\lambda)}{1-\Phi(\lambda)} - \left( \frac{\phi(\lambda)}{1-\Phi(\lambda)} \right)^2 \right) \right) + \\ (1 - \pi_i) \left( \left( \mu_i^{(i-1)*} - \sqrt{\Omega_{i,i}^{(i-1)*}} \frac{\phi(\lambda)}{\Phi(\lambda)} \right)^2 + \Omega_{i,i}^{(i-1)*} \left( 1 - \lambda \frac{\phi(\lambda)}{\Phi(\lambda)} - \left( \frac{\phi(\lambda)}{\Phi(\lambda)} \right)^2 \right) \right) - \\ \left( \pi_i (\mu_i^{(i-1)*} + \sqrt{\Omega_{i,i}^{(i-1)*}} \frac{\phi(\lambda)}{1-\Phi(\lambda)}) + (1 - \pi_i)(\mu_i^{(i-1)*} - \sqrt{\Omega_{i,i}^{(i-1)*}} \frac{\phi(\lambda)}{\Phi(\lambda)}) \right)^2 \quad \text{(Eq.S5)}$$

**Expected liability conditional on disease status of relatives**

To obtain the conditional expectation and variance of the other liabilities, we use the Pearson-Aitken selection formula, which says that if, conditioning on $Y_i$ and $K_i$ changes the expected mean liabilities from $\mu_i^{(i-1)*} = E(L_i | Y_1,..., Y_{i-1}, K_1, ..., K_{i-1}, K_{pop}, \Sigma)$ to $\mu_i^{i*} = E(L_i | Y_1, ..., Y_i, K_1, ..., K_i, K_{pop}, \Sigma)$, it will change the vector of liabilities μ to μ* , such that:

$$\mu^{i*} = \mu^{(i-1)*} + \Omega_{,i}^{(i-1)*} \left( \Omega_{i,i}^{(i-1)*} \right)^{-1} (\mu_i^{i*} - \mu_i^{(i-1)*}) \quad \text{(Eq.S6)}$$

Further if conditioning on $Y_n$ and $K_n$ changes the variance of liabilities from $\Omega_{i,i}^{(i-1)*} = \mathrm{Var}(L_i | Y_1,..., Y_{i-1}, K_1, \dots , K_{i-1}, K_{pop}, \Sigma)$ to $\Omega_{i,i}^{i*} = \mathrm{Var}(L_i | Y_1,..., Y_i, K_1, \dots K_i, K_{pop}, \Sigma)$, it will change the covariance matrix, Ω, to

$$\Omega^{i*} = \Omega^{(i-1)*} - \Omega_{,i}^{(i-1)*} \left( \left( \Omega_{i,i}^{(i-1)*} \right)^{-1} - \left( \Omega_{i,i}^{(i-1)*} \right)^{-1} \Omega_{i,i}^{i*} \left( \Omega_{i,i}^{(i-1)*} \right)^{-1} \right) \Omega_{i,}^{(i-1)*} \quad \text{(Eq.S7)}$$

**Example**

For an individual (3) with two family members (1 and 2) of which one is affected ($Y_1 = 1$) and the other is unaffected ($Y_2 = 0$) we can calculate the expected liability of individual 3, $E(G_3|Y_1 = 1, Y_2 = 0, K_1, K_2, K_{pop}, \Sigma)$, by the following procedure:
We start by setting the vector of liabilities,

$$\mu^{0*} = [E(L_1), E(L_2), E(G_3)]^T = [0, 0, 0]^T$$

with covariance matrix

$$\Omega^{0*} = \Sigma = \begin{bmatrix} Var(L_1) & Cov(L_1, L_2) & Cov(L_1, G_3) \\ Cov(L_2, L_1) & Var(L_2) & Cov(L_2, G_3) \\ Cov(G_3, L_1) & Cov(G_3, L_2) & Var(G_3) \end{bmatrix} = \begin{bmatrix} 1 & r_{1,2}h^2 & r_{1,3}h^2 \\ r_{2,1}h^2 & 1 & r_{2,3}h^2 \\ r_{3,1}h^2 & r_{3,2}h^2 & h^2 \end{bmatrix}$$

Next, we obtain the expected liability of relative 1, conditional on that individual being a case, (Eq.S4):

$$\mu_1^{1*} = E(L_1|Y_1 = 1, K_1, K_{pop}, \Sigma) = \pi_1(\mu_1^{0*} + \sqrt{\Omega_{1,1}^{0*}}\frac{\phi(\lambda)}{1-\Phi(\lambda)}) + (1 - \pi_1)(\mu_1^{0*} - \sqrt{\Omega_{1,1}^{0*}}\frac{\phi(\lambda)}{\Phi(\lambda)})$$

where since $\lambda = \frac{T-\mu_1^{0*}}{\sqrt{\Omega_{1,1}}} = T$ and $\pi_1 = P(D_1 = 1|Y_1 = 1, K_1, K_{pop}, \Sigma) = 1$, this simplifies to:

$$E(L_1|Y_1 = 1, K_1, K_{pop}, \Sigma) = \frac{\phi(\lambda)}{1-\Phi(\lambda)} = \frac{\phi(T)}{1-\Phi(T)}$$

And the expected variance:

$$\mathrm{Var}(L_1|Y_1, K_1, K_{pop}, \Sigma) \approx \pi_1\left(\left(\mu_1^{0*} + \sqrt{\Omega_{1,1}^{0*}}\frac{\phi(\lambda)}{1-\Phi(\lambda)}\right)^2 + \Omega_{1,1}^{0*}\left(1 + \lambda\frac{\phi(\lambda)}{1-\Phi(\lambda)} - \left(\frac{\phi(\lambda)}{1-\Phi(\lambda)}\right)^2\right)\right) +$$

$$(1 - \pi_1)\left(\left(\mu_1^{0*} - \sqrt{\Omega_{1,1}^{0*}}\frac{\phi(\lambda)}{\Phi(\lambda)}\right)^2 + \Omega_{1,1}^{0*}\left(1 - \lambda\frac{\phi(\lambda)}{\Phi(\lambda)} - \left(\frac{\phi(\lambda)}{\Phi(\lambda)}\right)^2\right)\right) -$$

$$\left(\pi_1(\mu_1^{0*} + \sqrt{\Omega_{1,1}^{0*}}\frac{\phi(\lambda)}{1-\Phi(\lambda)}) + (1 - \pi_1)(\mu_1^{0*} - \sqrt{\Omega_{1,1}^{0*}}\frac{\phi(\lambda)}{\Phi(\lambda)})\right)^2 =$$

$$\left(\left(\frac{\phi(T)}{1-\Phi(T)}\right)^2 + \left(1 + T\left(\frac{\phi(T)}{1-\Phi(T)}\right) - \left(\frac{\phi(T)}{1-\Phi(T)}\right)^2\right)\right) - \left(\frac{\phi(T)}{1-\Phi(T)}\right)^2 =$$

$$1 + T\left(\frac{\phi(T)}{1-\Phi(T)}\right) - \left(\frac{\phi(T)}{1-\Phi(T)}\right)^2$$

Now we update the vector of liabilities (Eq.S6):

$$\mu^{1*} = \mu^{0*} + \Sigma_{,1}\Sigma_{1,1}^{-1}(\mu_1^{1*} - \mu_1^{0*}) = [\,0, 0, 0]^T \; + \Sigma_{,1}\Sigma_{1,1}^{-1}\frac{\phi(T)}{1-\Phi(T)} =$$

$$\Sigma_{,1}\frac{\phi(T)}{1-\Phi(T)}$$

And the covariance matrix (Eq.S7):

$$\Omega^{1*} = \Omega^{0*} - \Omega_{,1}^{0*}\left(\left(\Omega_{1,1}^{0*}\right)^{-1} - \left(\Omega_{1,1}^{0*}\right)^{-1}\Omega_{1,1}^{1*}\left(\Omega_{1,1}^{0*}\right)^{-1}\right)\Omega_{1,}^{0*} = \Sigma - \Sigma_{,1}\left(\left(\frac{\phi(T)}{1-\Phi(T)}\right)^2 - \frac{T\phi(T)}{1-\Phi(T)}\right)\Sigma_{1,}$$

Having obtained the estimated liability conditional on the status of the first relative, we now condition on the status of the second relative, to estimate $\mu^{2*}$ and $\Omega^{2*}$.

First, the expected liability of relative 2 is:

$$\mu_2^{2*} = E(L_2|Y_1 = 1, Y_2 = 0, K_1, K_2, K_{pop}, \Sigma) = \pi_2\left(\mu_2^{1*} + \sqrt{\Omega_{2,2}^{1*}}\frac{\phi(\lambda)}{1-\Phi(\lambda)}\right) + (1 - \pi_2)\left(\mu_2^{1*} - \sqrt{\Omega_{2,2}^{1*}}\frac{\phi(\lambda)}{\Phi(\lambda)}\right)$$

where $\lambda = \frac{T-\mu_2^{1*}}{\sqrt{\Omega_{2,2}^{1*}}}$ and $\pi_2 = P(D_2 = 1\,|Y_1 = 1, Y_2 = 0, K_1, K_2, K_{pop}, \Sigma)$ which is obtained by (Eq.S3):

$$\pi_2 = 1 - \frac{\Phi\left(\frac{T-\mu_2^{1*}}{\sqrt{\Omega_{2,2}^{1*}}}\right)}{\Phi\left(\frac{T-\mu_2^{1*}}{\sqrt{\Omega_{2,2}^{1*}}}\right) + \frac{K_{pop}-K_2}{K_{pop}}\left(1-\Phi\left(\frac{T-\mu_2^{1*}}{\sqrt{\Omega_{2,2}^{1*}}}\right)\right)}$$

the conditional variance of the liability of relative 2 is:

$$\Omega_{2,2}^{2*} = \mathrm{Var}(L_2|Y_1 = 1, Y_2 = 0, K_1, K_2, K_{pop}, \Sigma) \approx \pi_2\left(\left(\mu_2^{1*} + \sqrt{\Omega_{2,2}^{1*}}\frac{\phi(\lambda)}{1-\Phi(\lambda)}\right)^2 + \Omega_{2,2}^{1*}\left(1 + \lambda\frac{\phi(\lambda)}{1-\Phi(\lambda)} - \left(\frac{\phi(\lambda)}{1-\Phi(\lambda)}\right)^2\right)\right)$$

$$(1 - \pi_2)\left(\left(\mu_2^{1*} - \sqrt{\Omega_{2,2}^{1*}}\frac{\phi(\lambda)}{\Phi(\lambda)}\right)^2 + \Omega_{2,2}^{1*}\left(1 - \lambda\frac{\phi(\lambda)}{\Phi(\lambda)} - \left(\frac{\phi(\lambda)}{\Phi(\lambda)}\right)^2\right)\right) -$$

$$\left(\pi_2\left(\mu_2^{1*} + \sqrt{\Omega_{2,2}^{1*}}\frac{\phi(\lambda)}{1-\Phi(\lambda)}\right) + (1 - \pi_2)\left(\mu_2^{1*} - \sqrt{\Omega_{2,2}^{1*}}\frac{\phi(\lambda)}{\Phi(\lambda)}\right)\right)^2$$

Again we update the vector of liabilities (Eq.S6):

$$\mu^{2*} = \mu^{1*} + \Omega_{,2}^{1*}\left(\Omega_{2,2}^{1*}\right)^{-1}(\mu_2^{2*} - \mu_2^{1*})$$

And the covariance matrix (Eq.S7):

$$\Omega^{2^*} = \Omega^{1^*} - \Omega^{1^*}_{,2}\left(\left(\Omega^{1^*}_{2,2}\right)^{-1} - \left(\Omega^{1^*}_{2,2}\right)^{-1}\Omega^{2^*}_{2,2}\left(\Omega^{1^*}_{2,2}\right)^{-1}\right)\Omega^{1^*}_{2,}$$

The third element of $\mu^{2^*}$ contains our estimate of $E(G_3|Y_1 = 1, Y_2 = 0, K_1, K_2, K_{pop}, \Sigma)$ i.e. the estimated liability of individual *3,* and the third diagonal element of $\Omega^{3^*}$ is its variance $\mathrm{Var}(G_3|Y_1 = 1, Y_2 = 0, K_1, K_2, K_{pop}, \Sigma)$ .
